# A dominant CIDEC variant segregates with familial obesity by increasing lipid droplet size in adipocytes

**DOI:** 10.1101/2025.07.28.25332029

**Authors:** Franziska Paul, Hui Wang, Jianqin Wang, Liang Juin Tan, Crystal Y. Chia, Lijuan Sun, Hui Jen Goh, Mark Kelly, Suresh Anand Sadananthan, Gunaseelan Narayanan, Lawrence M. Lifshitz, S. Sendhil Velan, Sarah Nicoloro, Sumanty Tohari, Alvin Y.J. Ng, Byrappa Venkatesh, Poh San Lai, Carine Bonnard, Kamille Airyel T. Enriquez-Satuito, Perie P. Adorable-Wagan, Elizabeth Paz-Pacheco, Eva Maria C. Cutiongco-De La Paz, Qijing Li, Melvin K. S. Leow, Li Xu, Li Peng, Michael P. Czech, Bruno Reversade

**Author notes:** **Corresponding Authors:** Franziska PAUL, Institute for Molecular and Cell Biology, A*STAR, 61 Biopolis Drive, Singapore 138673, Li PENG, School of Life Sciences, Tsinghua University, and Tsinghua-Peking Center for Life Sciences, Beijing, China, Michael P. CZECH, Program in Molecular Medicine, University of Massachusetts Chan Medical School, 366 Plantation Street, Worcester, MA 01605, Bruno REVERSADE, Laboratory of Human Genetics & Therapeutics, Biological and Environmental Sciences and Engineering Division (BESE), King Abdullah University of Science and Technology (KAUST), Thuwal, Kingdom of Saudi Arabia. These authors contributed equally to this article (Franziska Paul, Hui Wang).

## Abstract

Adipose tissue dysfunction in obesity is a major global public health risk, contributing to insulin resistance and chronic diseases such as diabetes and cardiovascular disorders. Here, we identify a dominant c.37A>G p.(Arg13Gly) variant in the long isoform of CIDEC (CIDEC-L), a key regulator of lipid droplet (LD) size, as the underlying cause of familial obesity. Affected individuals display marked subcutaneous fat accumulation in white adipose tissue (WAT), elevated fat content in brown adipose tissue (BAT) and insulin resistance. Accordingly, patient-derived iPSCs differentiated into white adipocytes exhibit accelerated LD growth, a phenotype mirrored by CIDEC-L^R13G^ overexpression. Mechanistically, we find that the p.Arg13Gly variant disrupts the N-terminal structural order of CIDEC-L, shifting its phase separation properties to enable, rather than restrict, lipid exchange through condensation plates between LDs. Notably, knock-in mice with the analogous Cidec-L p.(Arg10Gly) mutation recapitulate the human BAT hypertrophy and exhibit impaired thermogenesis. These findings establish the CIDEC-L^R13G^ variant as the first example of a dominantly inherited monogenic obesity driven by a dysfunctional adipocyte LD protein, revealing a critical role for CIDEC-L in restraining fat accumulation and maintaining metabolic health.

## Introduction

Highly synchronized coordination of lipid metabolism in adipose tissues is crucial for maintaining lipid storage, thermoregulation and energy homeostasis (Guilherme et al. 2008; Unger et al. 2010; Czech et al. 2013; Chen et al. 2020). Disruptions to this delicate balance, whether from excessive or insufficient lipid storage, or from gain or loss of adipose tissue itself, can lead to severe metabolic disorders (Czech 2017). The excessive white adipose tissue (WAT) accumulation in obesity, particularly in human visceral fat (fat around internal organs), is strongly associated with insulin resistance, liver steatosis, type 2 diabetes and cardiovascular disease (Lee et al. 2013; Czech 2020). On the other hand, a deficiency of adipose tissue, as observed in lipodystrophies, can also cause similar metabolic complications and other aspects of metabolic syndromes (Rubio-Cabezas et al. 2009; Brown et al. 2016; Zammouri et al. 2022). Altogether, this led to the general concept that a major beneficial function of metabolically healthy adipose tissue in lean - and even metabolically healthy obese - individuals, is to possess sufficient fat capacitance or adipose tissue expandability (Johannsen et al. 2014; Torres-Perez et al. 2015; Berger and Géloën 2023) via adipocyte hypertrophy and hyperplasia so as to sequester lipid away from other metabolic tissues such as liver and skeletal muscle, which are sensitive to toxic effects of lipid overload (Karpe and Pinnick 2015; Moreno-Indias and Tinahones 2015; Vishvanath and Gupta 2019). Accordingly, obesity that causes systemic metabolic dysfunctions is mostly associated with large lipid-laden white adipocytes in the visceral tissue that have lost the ability to sequester additional lipids. Such large adipocytes are also associated with secondary disruptions in adipose tissues, including defects in vascular function and increased inflammation (Reilly and Saltiel 2017). As a corollary, the dramatic improvement in insulin sensitivity among obese diabetic patients treated with thiazolidinediones (a class of PPARγ receptor agonists) has been attributed to fat remodeling associated with a shift from large unilocular adipocytes to smaller adipocytes with multilocular lipid droplets (LDs) with pronounced increased fat expandability. These beneficial changes are believed to result from expression of genes related to lipid metabolism and adipocyte differentiation (Toseland et al. 2001; Koh et al. 2009).

Adipocyte fat is stored in specialized organelles known as lipid droplets (LDs), which have a multitude of associated proteins on their surface controlling their size and ability to retain triglycerides (Puri and Czech 2008; Sztalryd and Brasaemle 2017; Olzmann and Carvalho 2019; Xu et al. 2024). LD size is determined by the relative rates of triglyceride hydrolysis (lipolysis), catalyzed by LD-associated lipases, versus triglyceride deposition and retention. One of the key drivers for LD size regulation is a family of three Cell death-inducing DNA fragmentation factor a–like effector (CIDE) proteins known as CIDEA, CIDEB and CIDEC (Puri et al. 2007; Keller et al. 2008; Puri et al. 2008; Slayton et al. 2019; Xu et al. 2024). CIDEB, which regulates LD size in the liver, was deemed a promising therapeutic target since rare germline loss-of-function variants were shown to protect from liver disease in a large exome sequencing cohort (Verweij et al. 2022). In contrast, CIDEA and CIDEC are predominantly regulating LD size in adipocytes. Two functional isoforms of CIDEC have been identified, a short form (CIDEC-S, FSP27α, NM_001321142, Uniprot:Q96AQ7-1) and a long form (CIDEC-L,

FSP27β, NM_001199623, Uniprot:A0A0A0MRY9), which are transcribed from different promoters and differ by 13 additional N-terminal amino acids in human, and 10 amino acids in mouse (Nishimoto et al. 2017; Nishimoto and Tamori 2017).

Previous findings in mice show that the ratio of Cidec-S and Cidec-L isoforms expressed in adipose tissue correlates with LD size, suggesting that they might have antagonistic roles (Nishimoto et al. 2017). A high proportion of Cidec-S promotes LD growth in white adipose tissue (WAT), while the predominant Cidec-L isoform restricts LD size in thermogenic, multilocular adipocytes from brown adipose tissue (BAT) (Toh et al. 2008; Nishimoto et al. 2017; Nishimoto and Tamori 2017). It is likely that this ratio of CIDEC isoforms can oscillate by increasing the long form expression relative to the short form during browning of WAT while reducing this long form and increasing the short form during whitening of BAT. Consistent with this notion, Cidec null mice lacking both isoforms show reduced LD size in WAT but increased LD size in BAT (Toh et al. 2008). Owing to the lack of Cidec-S in WAT, these Cidec null mice failed to efficiently store triglyceride in their reduced subcutaneous and gonadal fat depots, thereby being protected from obesity (Nishino et al. 2008; Zhou et al. 2015). However, the defective lipid storage in WAT dramatically elevated circulating triglycerides, which, as a result of concomitant Cidec-L deficiency, led to ectopic fat accumulation in both BAT and liver, resulting in hepatic steatosis and insulin resistance (Zhou et al. 2015). Consistent with these findings in rodents, a homozygous stop-gained allele exhibiting autosomal recessive inheritance in a human subject causing a combined loss-of-function of both CIDEC-S and CIDEC-L proteins resulted in insulin-resistant diabetes and severe fat atrophy (Rubio-Cabezas et al. 2009).

Here we report on an extended kindred with a heterozygous germline c.37A>G; p.(Arg13Gly) variant that exclusively affects CIDEC-L, leaving CIDEC-S intact. Nine individuals over three generations presented with striking obesity which rapidly worsened with age. Patient-derived iPSCs differentiated into WAT displayed accelerated LD growth, and knock-in mice with the homologous Cidec-L^R10G^ mutation developed enlarged LD in BAT leading to impaired thermogenesis. In cell lines, we document that CIDEC-L normally functions to restrict CIDEC-S-driven lipid exchange between droplets. Mechanistically, this can be attributed to differential N-terminal phase separation properties of both isoforms. We find that the patient variant CIDEC-L^R13G^ displays phase separation properties more closely resembling CIDEC-S, thus losing its ability to restrict LD growth. This lack of LD size restriction likely accounts for the excessive fat accumulation in both WAT and BAT of CIDEC-L^R13G^ carrier patients, in addition to its increased intrinsic activity.

## Results

### A CIDEC-L^R13G^ variant segregates with familial obesity

We investigated an extended family presenting with hereditary obesity which manifested as progressive, symmetrical enlargement of the trunk and the proximal areas of both upper and lower extremities, suggestive of WAT accumulation. In the index case (I:1), this striking phenotype became progressively more conspicuous in his late forties without being related to increased physical activity or diet change (Fig. 1A-B and Table 1). Similar symptoms were recorded in several of his children and grandchildren, irrespective of gender, suggestive of an autosomal dominant inheritance (Fig. 1A-B and Table 1). A post-mortem biopsy of the deceased index patient I:1 and an MRI of his daughter’s (II:4) leg, indicated that the enlargement of the proximal extremities was due to excessive accumulation of subcutaneous white adipose tissue (Fig. 1C-D). To identify the suspected underlying genetic etiology of the disease, we performed trio exome sequencing of individuals I:2, II:4 and II:8. Candidate gene identification relied on the filtering of germline variants that were autosomal, heterozygous, coding, rare (Minor Allele Frequency: MAF<0.001) and predicted to be pathogenic (Combined Annotation Dependent Depletion: CADD>20). Only one private variant c.37A>G; p.(Arg13Gly) in the long isoform (NM_001199623) of the *CIDEC* gene (“CIDEC-L”, Fig. 1E) segregated with the disease phenotype in affected individuals, while available non-affected family members were non-carriers (Fig. 1A). This variant is observed only once in the heterozygous state in the gnomAD database (v4.1.0), which collects variants from the general population, and has never been observed in the homozygous state. It is not reported in ClinVar, a freely accessible database of reports of human variations classified for diseases and drug responses. The evolutionary conservation of the Arginine 13 residue in CIDEC-L across vertebrates (Fig. 1F) underscores its functional importance, while its substitution with Glycine is predicted to be deleterious with a CADD score of 26.2 (Fig. 1G).

**Figure 1.**
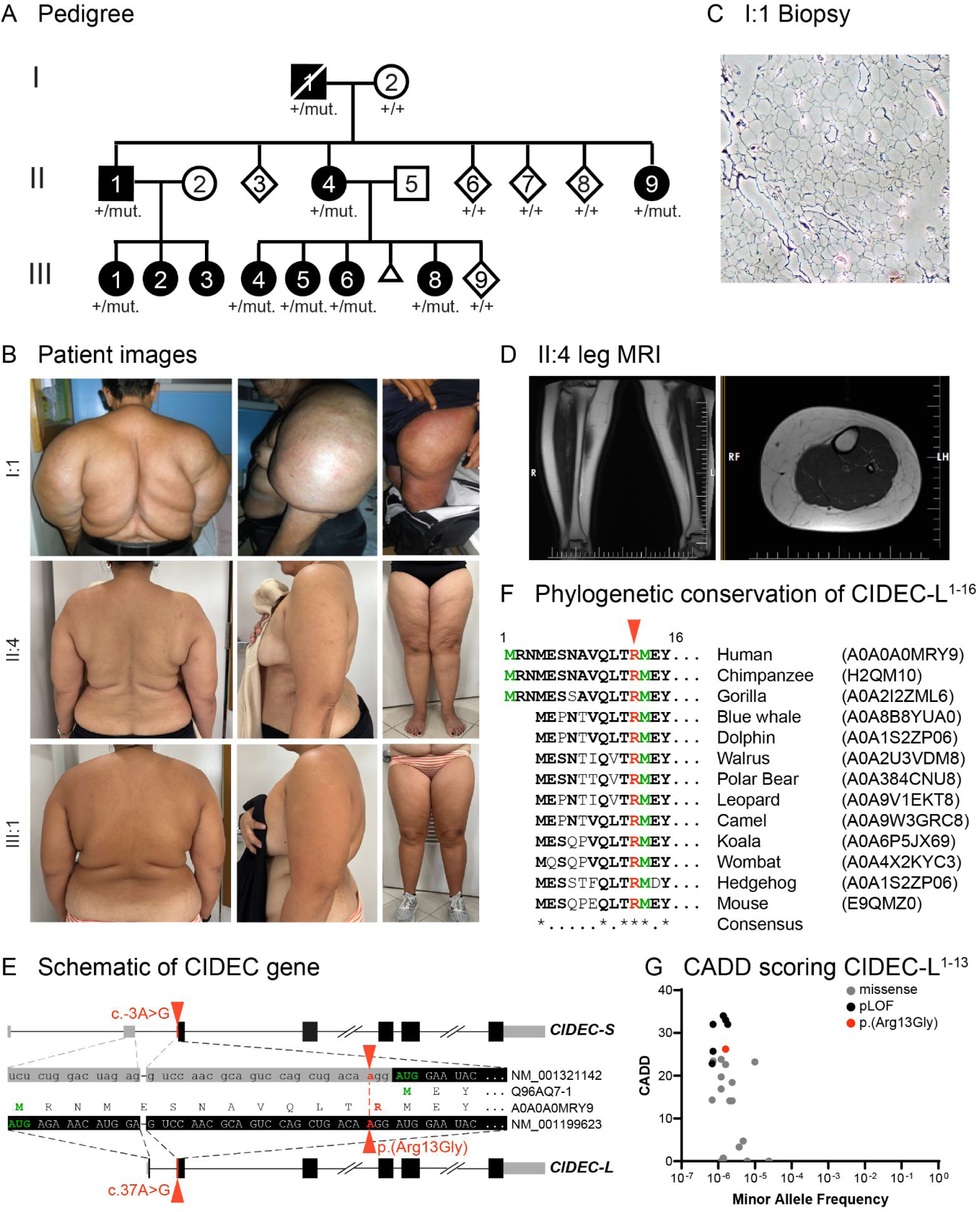
A dominantly inherited form of obesity segregates with the CIDEC-L^R13G^ missense variant. **Panel A** shows the pedigree of a family with progressive fat accumulation across generations. Squares indicate males, circles females, open symbols unaffected, black symbols affected and diagonal slash deceased. After whole exome sequencing, the mutant gene was identified: ‘+’ denotes wildtype, and ‘mut.’ for mutant alleles. **Panel B** shows images of the back, upper arm and thighs from affected individuals including the proband (I:1), his daughter (II:4) and a granddaughter (III:1). **Panel C** depicts a puncture biopsy taken from the I:1 proband’s thigh, revealing the unilocular adipocytes structures in the affected tissues. **Panel D** shows X-ray images of the proband’s daughter II:4, highlighting the accumulation of fat subcutaneously. **Panel E** shows a schematic of the germline mutation c.37A>G; p.(Arg13Gly) missense variant specific to the long isoform of CIDEC. **Panel F** depicts the strict phylogenetic conservation of the CIDEC-L^R13^ residue across evolution. **Panel G** shows the Minor allele frequency (MAF) against the combined annotation-dependent depletion (CADD) score for coding variants found in the first 13 residues of CIDEC-L as listed in gnomAD v.4.0.0 (black and grey dots) and CIDEC-L^R13G^ found in our family (orange). Notably CIDEC-L^R13G^ clusters with loss of function variants.

**Table 1.**
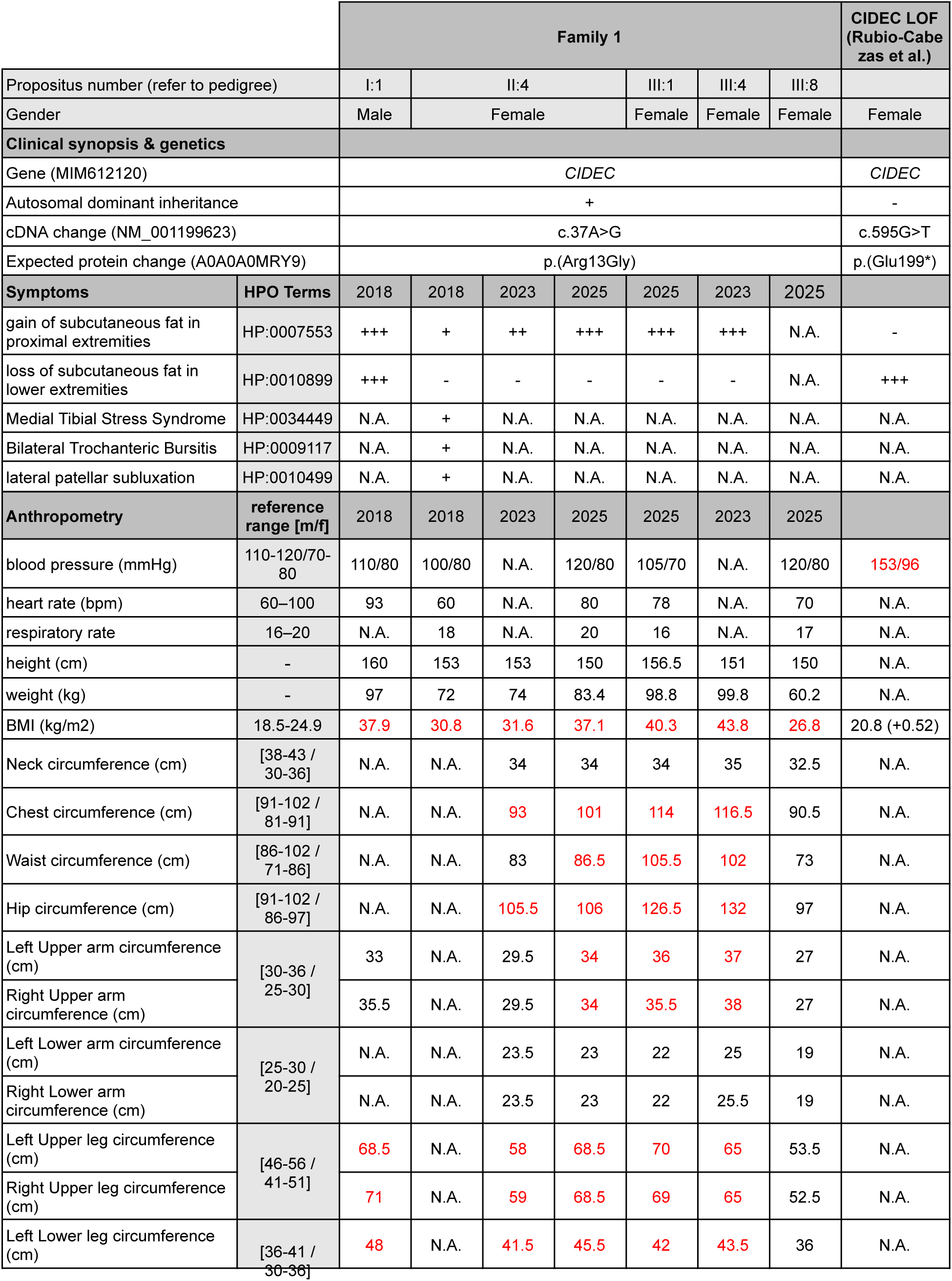
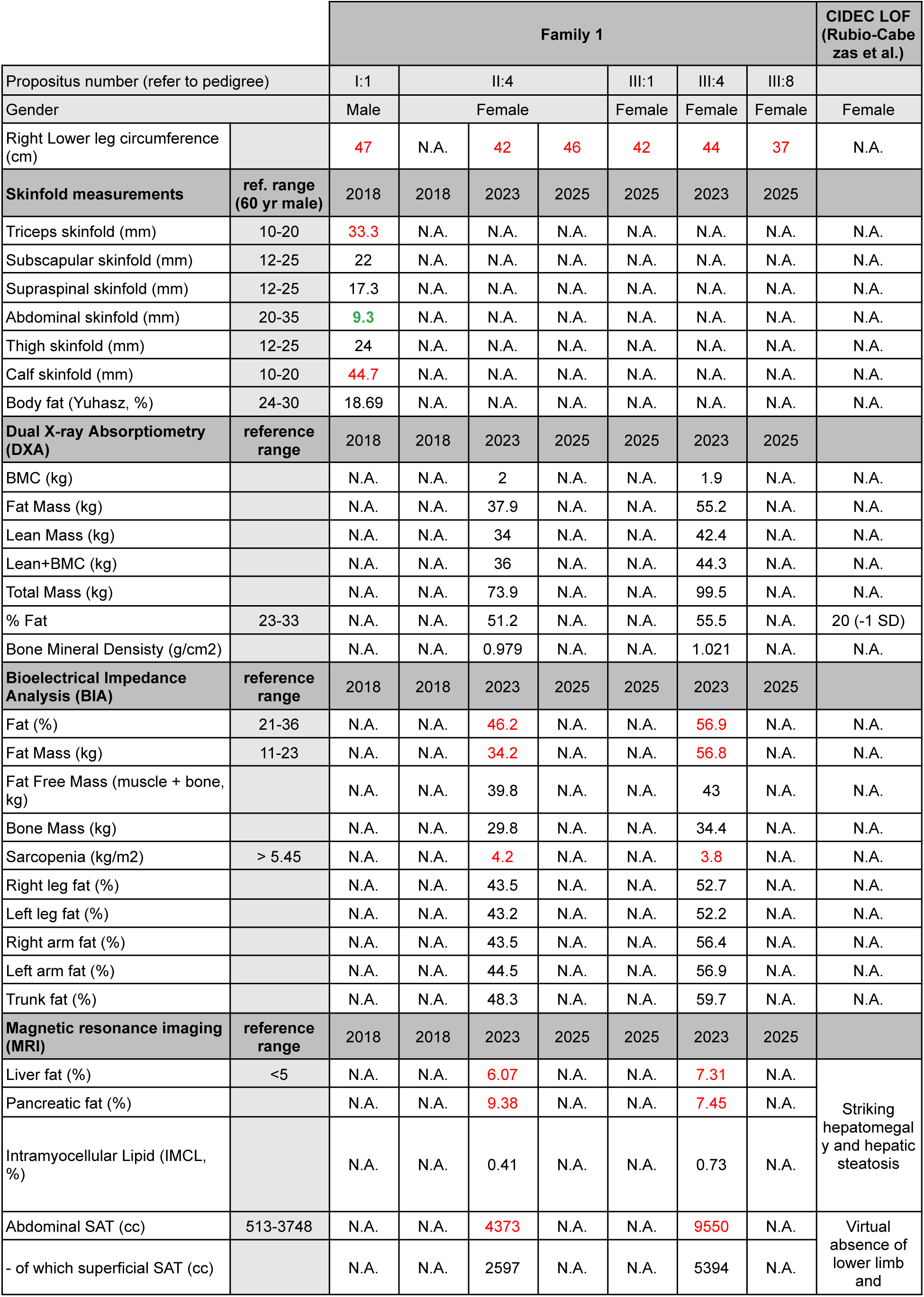
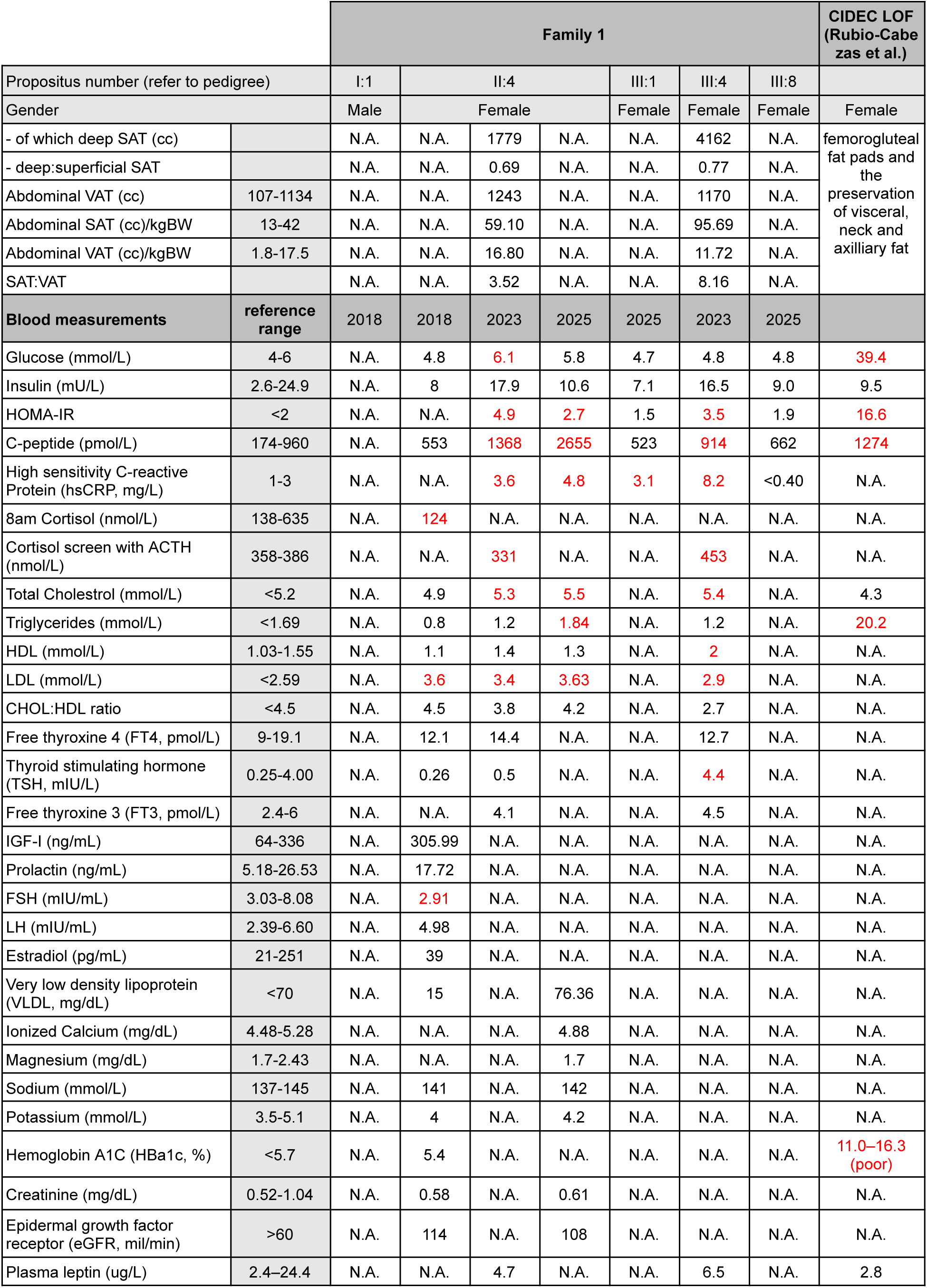
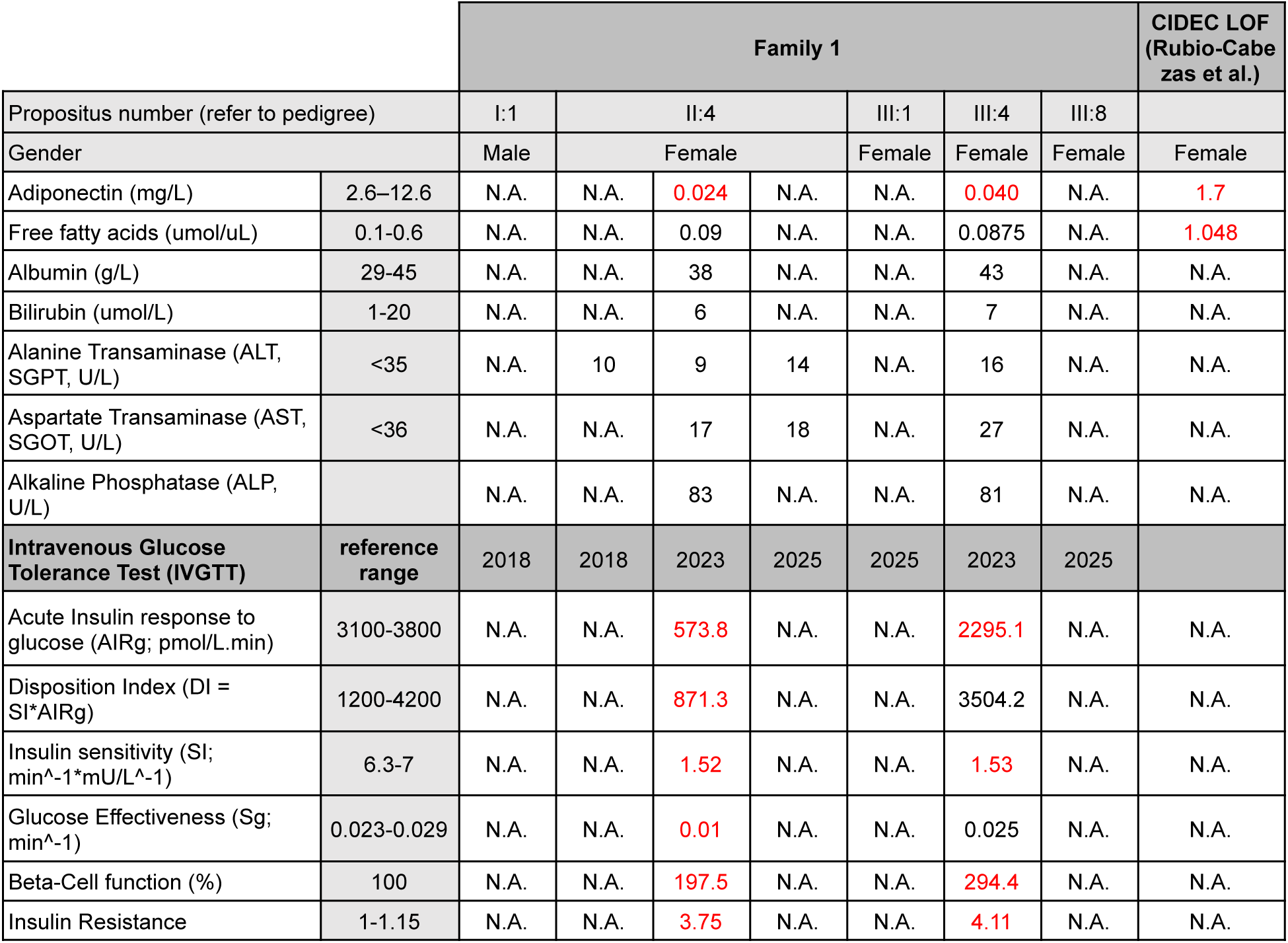
Clinical characteristics of patients with pathogenic germline CIDEC variants.

### CIDEC-L^R13G^ individuals display enlargement of adipose tissue and insulin resistance

We subsequently invited two carriers of the CIDEC-L^R13G^ variant for deeper phenotyping, designed to assess their adiposity and metabolic profile. Both females, a mother (II:4) and her daughter (III:4), exhibited an elevated BMI (37.1 and 43.8, respectively) consistent with marked obesity. A dual-energy X-ray absorptiometry (DXA) scan illustrated the extensive accumulation of white fat in the abdomen, pelvis, back, gluteal area and proximal extremities (Fig. 2A), adding up to a total body fat of 51.2% and 55.5%, respectively (Table 1). While their basal plasma glucose and insulin levels were on the higher end of the normal range (Table 1), an intravenously administered glucose tolerance test (IVGTT) revealed markedly reduced insulin sensitivity in both individuals, with one individual manifesting impaired fasting glycemia or pre-diabetes (subject II:4), indicating the development of insulin resistance (Table 1). Concurrent with findings in mice (Nishimoto et al. 2017), *CIDEC-L* is the predominant isoform expressed in BAT of non-human primate *Macaca fascicularis* (Fig. 2B). Thus, we examined the supraclavicular BAT depots of patients II:4 and III:4 by 18-fluorine fluorodeoxyglucose positron emission tomography (18F-FDG PET) scan, which are known to contain thermogenic, multilocular adipocytes (Wu et al. 2012; Ikeda et al. 2018). The patients’ BAT depots remained functionally active as evidenced by the increased 18F-FDG uptake after cold stimulation (Fig. 2C). Interestingly, we detected a highly elevated fat content (“fat fraction”, FF) of ∼85-92% in these adipose depots that more closely resembles the fat content of white adipocytes (reference values from other in-house studies show FF<70% for human BAT/“beige” adipose depots and FF≥90% for WAT, Fig. 2D). This suggests that the supraclavicular adipocytes in these patients have larger LD to accommodate a high quantity of triglycerides. Notably, the FF value reduced after cooling (Fig. 2D), suggestive of cold-induced lipolysis that is typical for thermogenic BAT. While CIDEC-L is the predominant isoform in BAT, it is also expressed in WAT (Nishimoto et al. 2017), which explains the dynamic nature of LD remodeling in WAT from unilocular white adipocytes to multilocular LD in beige adipocytes when WAT undergoes browning. In line with this, the CIDEC-L^R13G^ variant likely causes larger unilocular LD and adipocyte hypertrophy, accounting for the excessive accumulation of subcutaneous fat in our patients.

**Figure 2.**
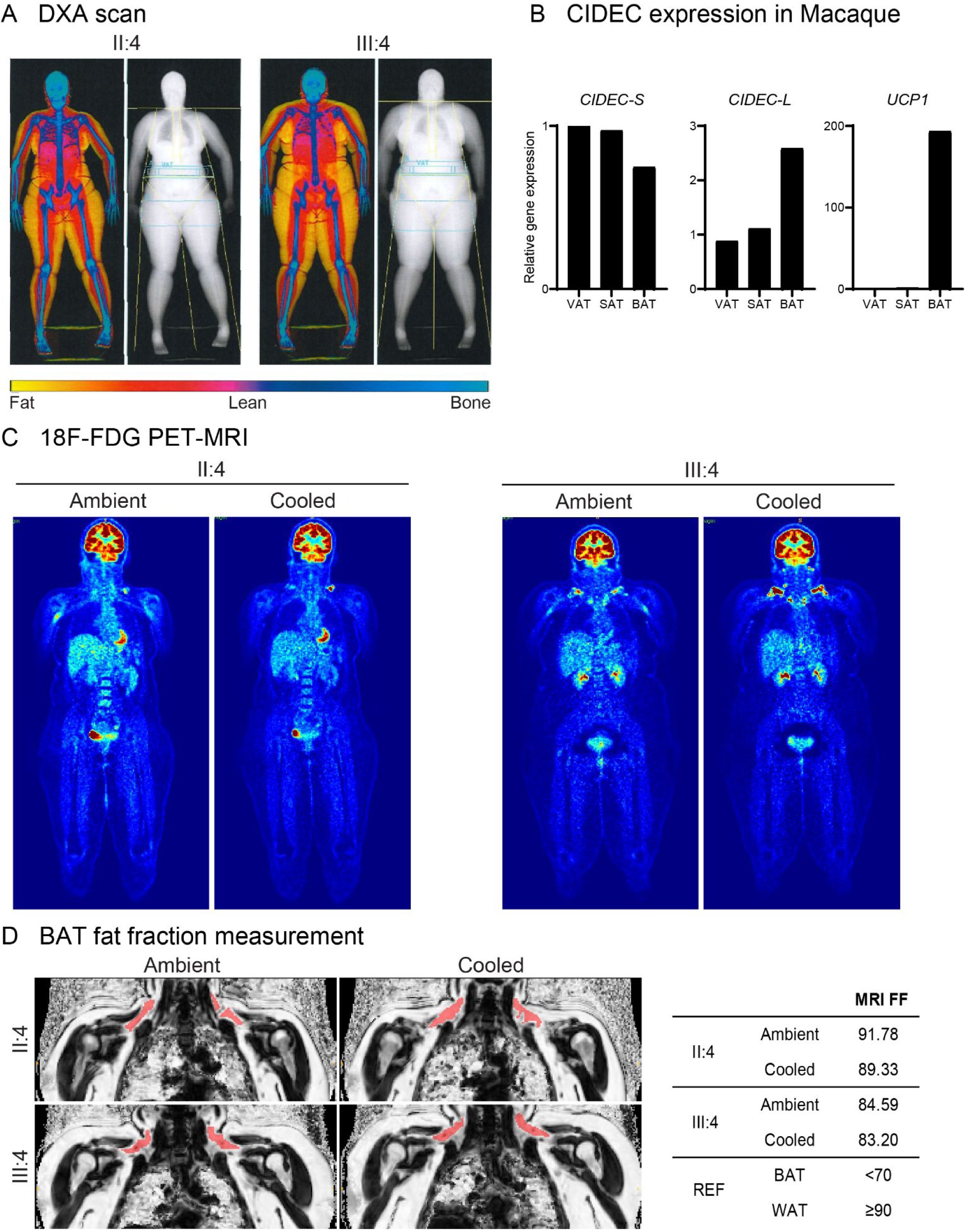
CIDEC-L^R13G^ mutation causes fat accumulation in humans. **Panel A** shows dual energy X-ray absorptiometry (DXA) scans of daughter II:4 and granddaughter III:4, which illustrates the extensive subcutaneous fat accumulation (shown as yellow color). **Panel B** shows RNA expression of the *Cidec* isoforms relative to *Rpl4* across fat tissues from one individual of *Macaca fascicularis*. The *Cidec-L* isoform is expressed at high levels in BAT and at lower levels in WAT. **Panel C** depicts positron-emission tomography and magnetic resonance imaging (PET-MRI) scans after injection of ^18^F-FDG. Images on the right were taken after wearing a cooling vest for two hours and confirm that the supraclavicular BAT depots are actively taking up glucose after cold stimulation. **Panel D** shows ^18^F-FDG-PET scan images of the upper thorax of two affected individuals under ambient temperatures and after wearing a cooling vest for two hours. The active human brown/beige adipose tissue (BAT) in the supraclavicular region is highlighted in red. Fat fraction (FF, a measure for fat content) was determined by MRI within the red BAT regions as summarized in the table on the right. In-house reference values (“REF”) for FF of BAT and WAT are indicated in the table.

### Patient-derived CIDEC-L^R13G^ white adipocytes display rapid LD expansion

To capture the dynamics of LD growth in CIDEC-L^R13G^ white adipocytes, we derived induced pluripotent stem cells (iPSCs) from patient II:4 and differentiated them into mesenchymal progenitor cells (MPCs), which were then matured into white adipocytes (Lee and Cowan 2014). Notably, patient-derived MPC exhibited accelerated LD formation, with visible droplets appearing as early as 3 days post-differentiation, compared to 1 week in control MPCs. At 14 days post-MPC differentiation, immunofluorescence staining revealed significantly larger LDs in patient-derived white adipocytes compared to control cells (Fig. 3A-B). The elevated expression of *ADIPOQ*, *FABP4*, *CEBPA* and *PPARG* confirmed the robust differentiation state of the adipocytes (Fig. 3C). At 21 days post-MPC differentiation, while both patient and control cells reached similar maximal droplet sizes, a striking absence of small LDs was documented in patient-derived adipocytes (Fig. 3D-E). These contrasting dynamics suggest that CIDEC-L^R13G^ fails to decelerate LD growth in cultured human white adipocytes. Our *in vitro* results are thus recapitulating the observed WAT enlargement seen in the affected individuals, indicating that CIDEC-L plays an instructing role in LD dynamics.

**Figure 3.**
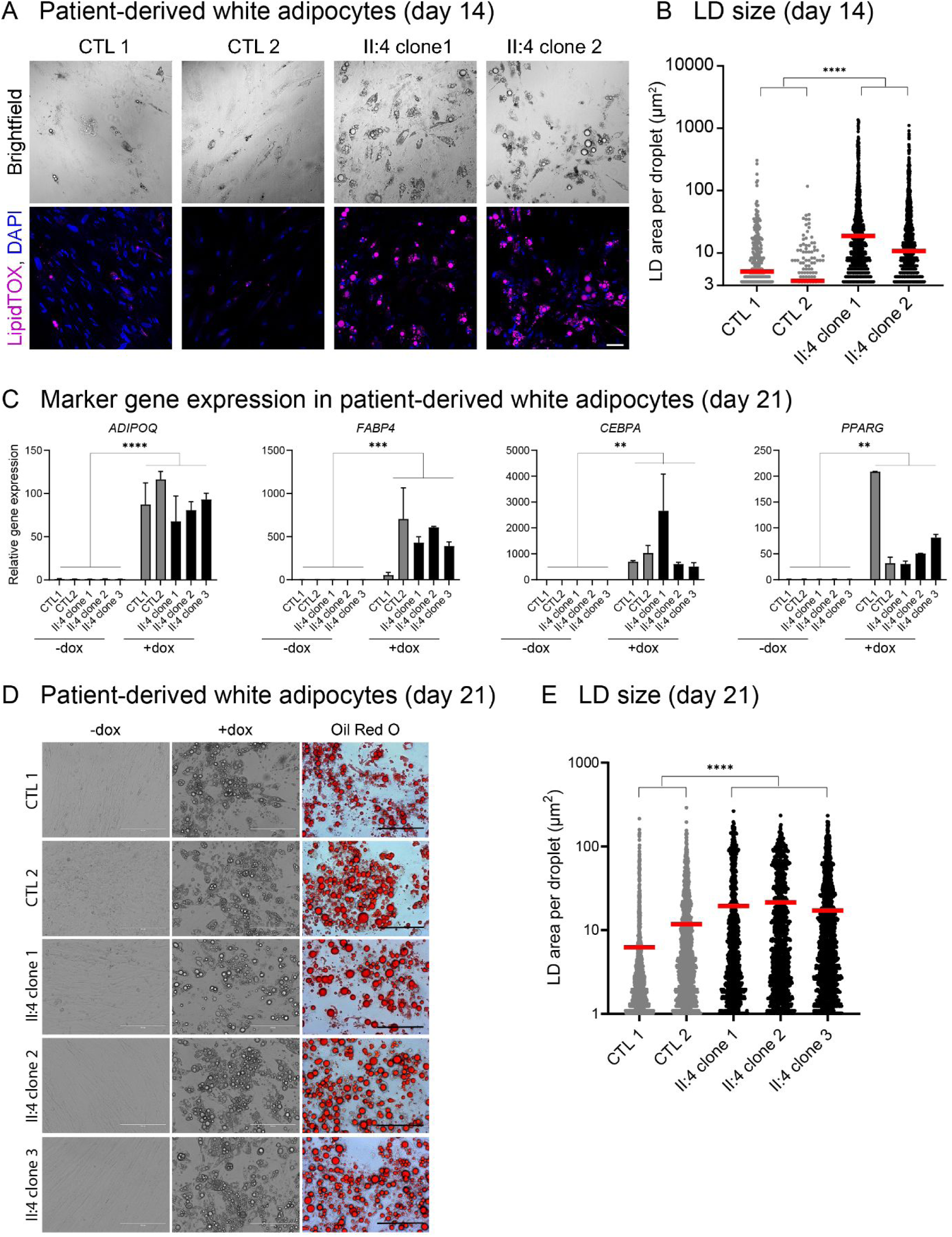
Lipid droplets grow faster in patient-derived white adipocytes. **Panel A** shows Patient-derived induced pluripotent stem cells (iPSC) that were differentiated into mesenchymal progenitor cells (MPC) and then into white adipocytes. In contrast to unrelated control cells, the CIDEC-L^+/R13G^ patient-derived MPC started forming visible droplets after only 3 days, which then grew proportionally larger. Immunofluorescence staining of differentiated white adipocytes at 14 days post MPC differentiation. LipidTOX stains LDs (pink) and DAPI nuclei (blue). Scale bar = 100 μm. **Panel B** LD size quantified from Panel A, which were measured with the ImageJ Particle analyzer (circularity 0.5-1, size 0-infinity). Two-tailed Mann-Whitney test for combined patient vs. control cells; **** *p*<0.0001. **Panel C** shows the gene expression profile of fully differentiated (day 21) white adipocytes derived from patient MPC. Gene expression levels were normalized to human housekeeping gene *HPRT1*, n=2 for each condition, normalized to no doxycycline, combined paired two-sided t-test: ** *p* <0.01, *** *p* <0.001, **** *p* <0.0001. **Panel D** Doxycycline induced the formation of LDs in patient-derived white adipocytes, which reached comparable maximum sizes after 21 days in culture. White scale bar = 200 μM, black scale bar = 100 um. **Panel E** shows the quantified LD size from patient-derived white adipocytes in Panel D. Lipid droplet size was measured with ImageJ particle analyzer: size cutoff >1 μm^2^, Mann-Whitney test *p*-value **** <0.0001.

### CIDEC-L^R13G^ lacks LD size restriction properties

Both CIDEC-S and CIDEC-L are recruited to the surface of lipid droplets where they antagonistically regulate lipid transfer between droplets (Nishimoto and Tamori 2017; Lyu et al. 2021). To examine the effects of CIDEC-S and CIDEC-L on LD size, they were transiently over-expressed in 3T3-L1 preadipocytes. Forced CIDEC-L^WT^ expression generated LDs that were 4 to 5 times smaller than those of CIDEC-S, while its mutant variant CIDEC-L^R13G^ was unable to limit LD growth (Fig. 4A). This may be explained by an elevated lipid exchange rate between LD (Fig. 4B), and an increase in LD-LD contact sites (LDCS, Fig. 4C). The localization of CIDEC-L^R13G^ to LDCS was unaffected (Fig. 4D).

**Figure 4.**
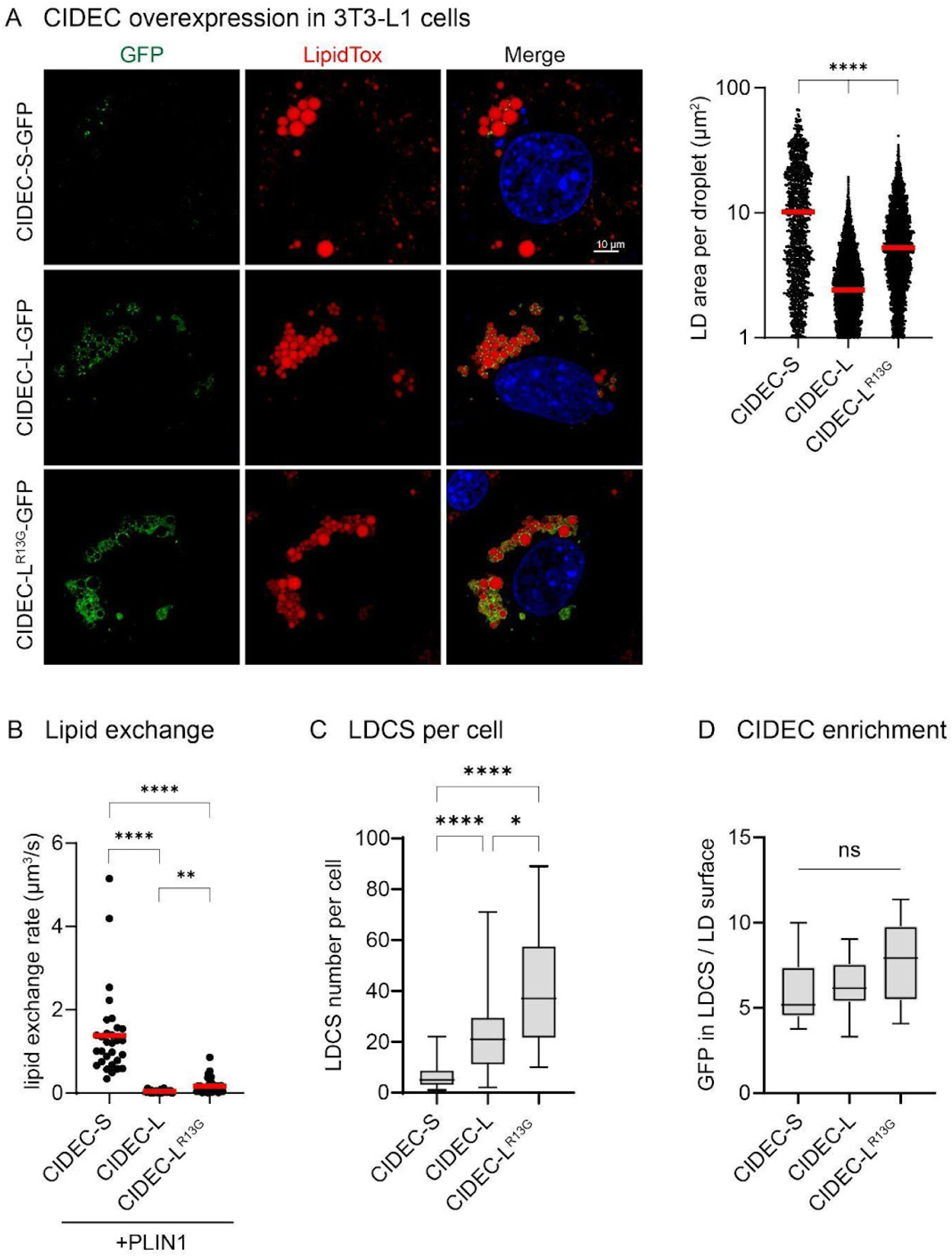
CIDEC-L^R13G^ promotes LD fusions in 3T3-L1 pre-adipocytes. **Panel A** shows the overexpression of GFP-tagged CIDEC variants in 3T3-L1 cells. Representative immunofluorescence images are depicted on the left, with LipidTox stained LDs (red). Introducing the p.R13G patient mutation into CIDEC-L causes an increase of LD size as quantified on the right (Kruskal-Wallis Test). This can likely be attributed to an increase of the lipid exchange rate (under co-transfection with *PLIN1*, unpaired two-tailed t-test, **Panel B**) and an increased number of lipid droplet contact sites per cell (LDCS, Kruskal-Wallis Test, **Panel C**). The localization of CIDEC-L to LDCS is not affected by the p.R13G variant (one-way ANOVA, p>0.05, **Panel D**). * *p* <0.05, ** *p* <0.01, *** *p* <0.001, **** *p* <0.0001 for all tests.

At the LDCS, it is thought that CIDEC oligomerizes with other CIDE proteins via its CIDE-N domain that lies downstream of the p.R13G missense mutation (Choi et al. 2017; Lyu et al. 2021; Xu et al. 2024). To investigate whether CIDEC-L^R13G^ retained the ability to form homomers, we performed co-immunoprecipitation in HEK293T cells. While the first 39 common residues are important for homomerization, the p.Arg13Gly mutation in CIDEC-L did not prevent this process (Fig. 5A). Moreover, CIDEC-L^R13G^ retained the ability to form heteromers with CIDEC-S and CIDEA (Fig. 5B), which are the predominant proteins driving LD growth in WAT and BAT, respectively (Wu et al. 2014; Barneda et al. 2015; Nishimoto et al. 2017; Nishimoto and Tamori 2017). Surprisingly, when co-expressing CIDEC isoforms in 3T3-L1 cells, we found that CIDEC-L showed an inhibitory effect on CIDEC-S-mediated LD growth, while the CIDEC-L^R13G^ variant partially lost its size-restriction effect on CIDEC-S (Fig. 6A). Similarly, CIDEC-L restricted CIDEA-mediated LD growth, which was partly abrogated when the CIDEC-L^R13G^ variant was introduced (Fig. 6B). Taken together, these data imply that CIDEC-L^R13G^ cannot perform its repressive role vis-à-vis CIDEC-S or CIDEA, and therefore loses its ability to curb LD size increase in both WAT and BAT.

**Figure 5.**
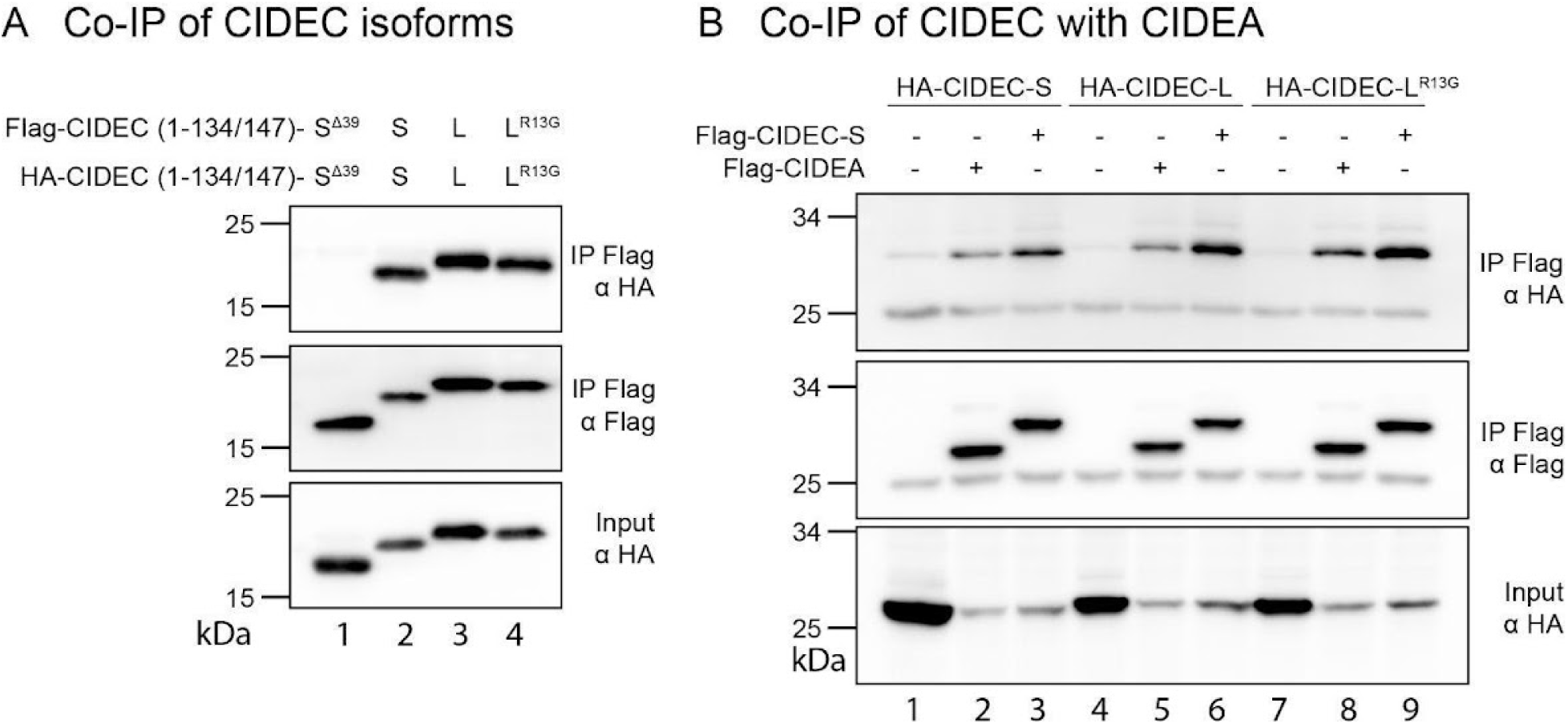
CIDEC-L^R13G^ does not impact binding to other CIDE proteins. We performed co-immunoprecipitation (Co-IP) of N-terminal CIDEC variants to demonstrate that CIDEC-S and CIDEC-L form homo-mers (**Panel A**). The homotypic binding of CIDEC-L is not impaired by the R13G mutation (lane 4). Moreover, CIDEC-L and CIDEC-L^R13G^ retain the ability to bind to CIDEC-S and CIDEA as shown by Co-IP in **panel B**.

**Figure 6.**
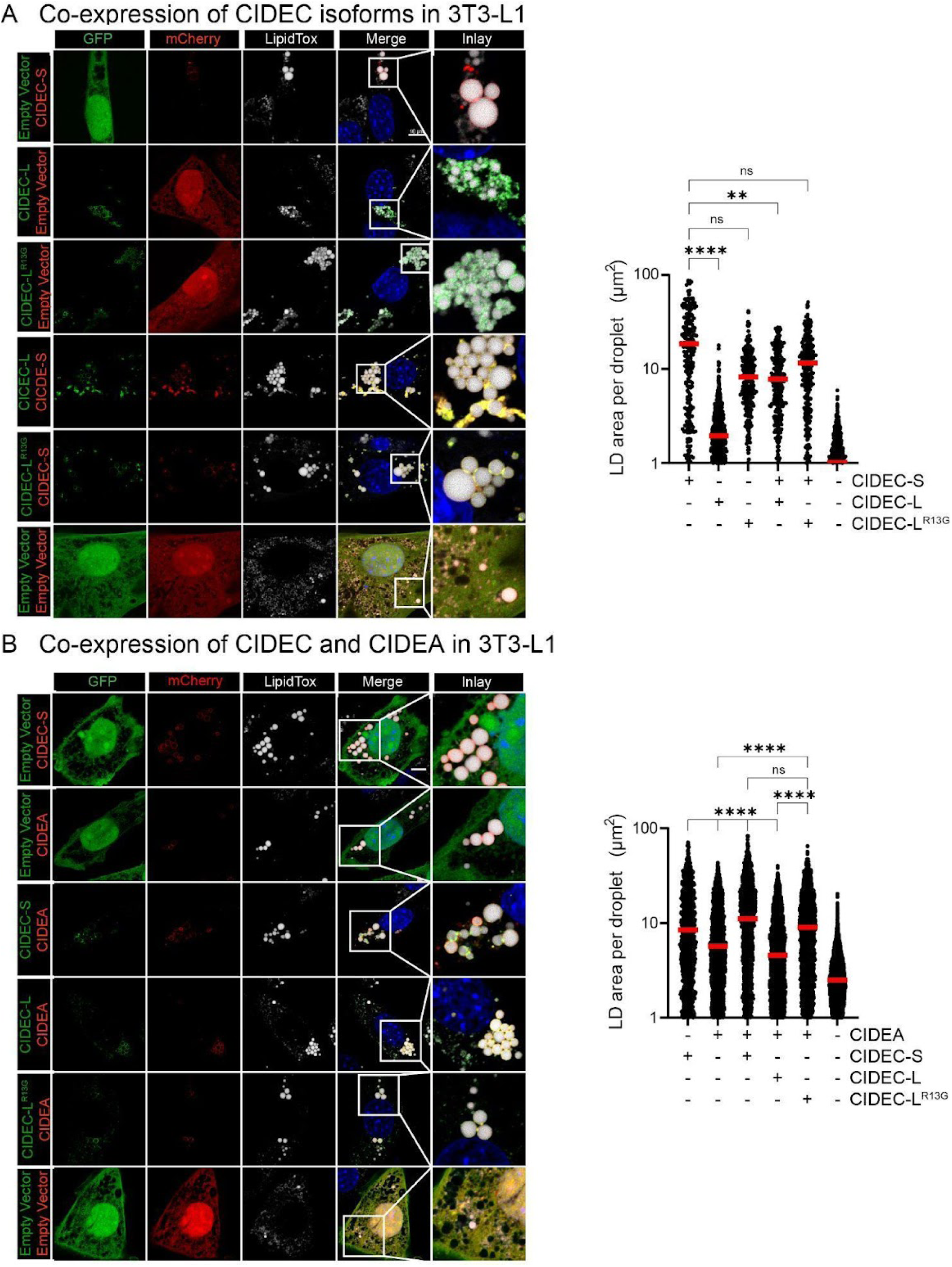
CIDEC-L^R13G^ displays less inhibitory effect on CIDEC-S or CIDEA-mediated LD growth. We co-expressed fluorescently tagged CIDEC isoforms in 3T3-L1 cells, showing that the p.R13G mutation does not impact CIDEC-L’s ability to co-localize with CIDEC-S (**Panel A**). LDs size quantifications are shown on the right. Importantly, CIDEC-L and CIDEC-L^R13G^, but not CIDEC-S, also co-localize with CIDEA when overexpressed in 3T3-L1 cells (**Panel B**). LDs size quantifications are also on the right.

Since the CIDEC-L^R13G^ variant did not display impaired localization nor oligomerization, we sought to model the behavior of its N-terminal tail using AlphaFold3 (Fig. 7A-B). Reiterative predictions suggested that the p.Arg13Gly mutation causes a loss of secondary structure in the N-terminus first 13 residues of CIDEC-L which in its WT sequence folds into a predicted alpha helix. Since structural disorder is associated with phase separation necessary for the formation and function of LDCS (Lyu et al. 2021), we performed a cell-free phase separation assay using purified CIDEC proteins (Fig. 7C). While CIDEC-S possessed strong phase separation properties, the additional 13 N-terminal residues of CIDEC-L hindered phase separation. Notably, introduction of the single amino acid change p.R13G made CIDEC-L^R13G^ behave like CIDEC-S, achieving phase separation. This intrinsic difference is likely to promote CIDEC-L^R13G^ condensation within CIDE complexes at the LDCS, and thus favor lipid transfer between droplets (Lyu et al. 2021). Together, these datasets establish a loss-of-function for the CIDEC-L^R13G^ variant which, unlike native CIDEC-L, is unable to repress CIDEC-S-mediated LD growth in both BAT and WAT. In the affected individuals we speculate that this manifests as elevated fat content in BAT and enlargement of WAT respectively.

**Figure 7.**
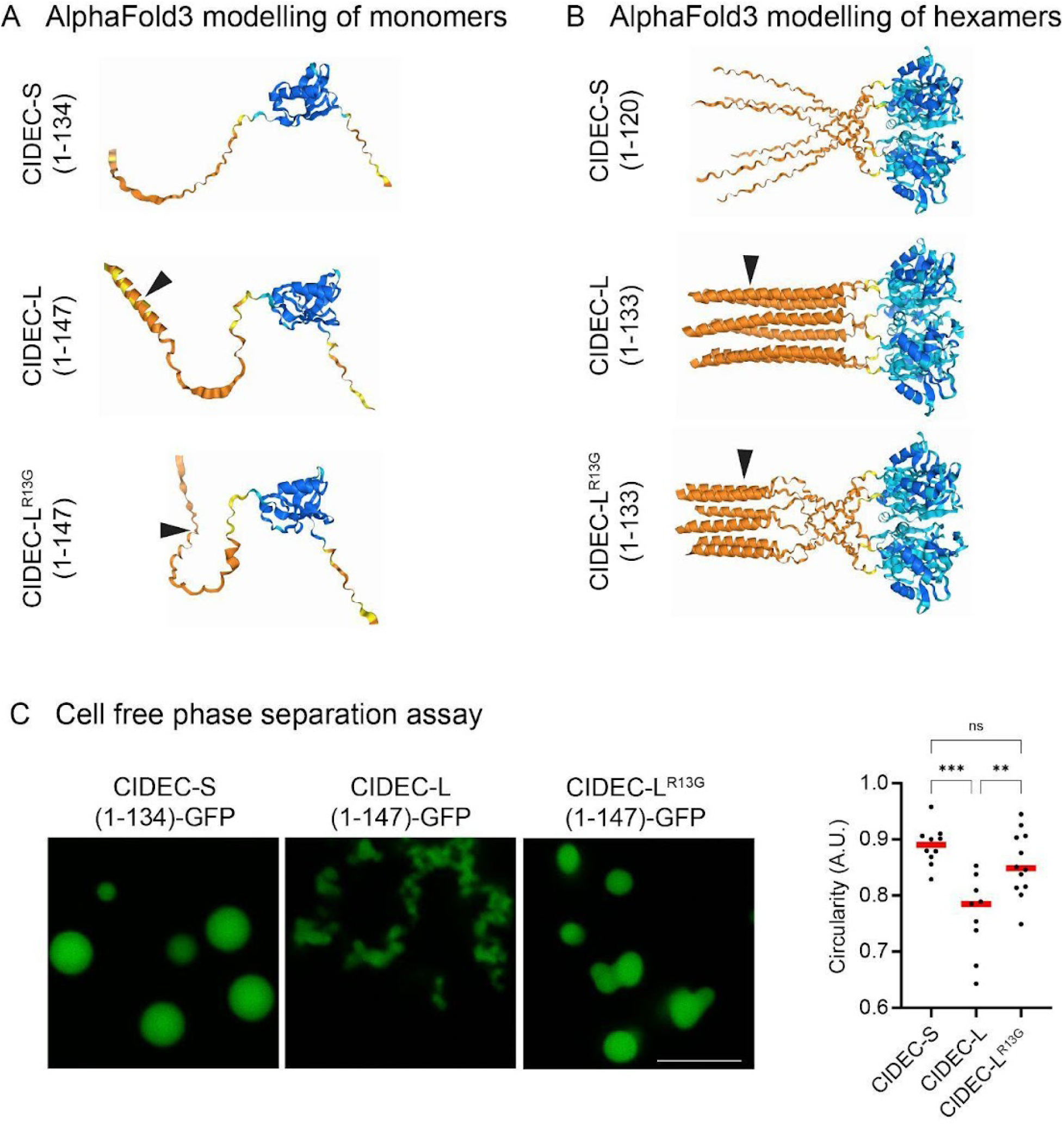
Structural N-terminal disorder in CIDEC-L^R13G^ is correlated with phase separation. Structural modelling of the N-terminal end of CIDEC monomers (residues 1-134 for CIDEC-S or 1-147 for CIDEC-L; **Panel A**) and hexamers (residues 1-120 for CIDEC-S or 1-133 for CIDEC-L; **Panel B**) using AlphaFold3. Confidence as follows: blue = Very high (plDDT > 90), teal = Confident (90 > plDDT > 70), yellow = Low (70 > plDDT > 50), orange = Very low (plDDT < 50). The black arrow marks residue 13 in CIDEC-L. Albeit the model of the N-terminal tail is built on low confidence, it is consistently modelled as an alpha helix in CIDEC-L, with the R13G variant breaking this structure. This structural disorder is correlated with condensation properties (**Panel C**). Phase separation assay was performed on 50 μM purified protein in 10% PEG 4000, 150 nM Nacl and 25 mM Tris. Circularity is quantified on the left (One-way ANOVA).

### Knock-in Cidec-L^R10G^ mice have enlarged LDs in BAT and fail to thermoregulate

To mimic a mutation analogous to the above patients’ variant, we established a knock-in mouse model incorporating the pathogenic missense (NM_001372264.1, c.28A>G, p.Arg10Gly of *Cidec-L*) into the mouse *Cidec* locus via CRISPR/Cas9-mediated genome editing (Fig. 8A). The genomic DNA sequences were validated through Sanger sequencing (Fig. 8B). Both heterozygous Cidec-L^R10G/+^ and homozygous Cidec-L^R10G/R10G^ knock-in mice displayed normal health and comparable viability as did wild-type Cidec-L^+/+^ controls. To more precisely recapitulate the dietary exposome associated with this human disease, all mice were subjected to a Western-style diet enriched in saturated fat, cholesterol, and sucrose to promote obesity and insulin resistance. Based on the higher level of *Cidec-L* expression in BAT and brown adipocytes (Fig. 8C-D) as noted previously (Nishimoto et al. 2017), we hypothesized that Cidec-L^R10G^ may interfere with the BAT function in non-shivering thermogenesis. Therefore, mice were subjected to a cold exposure regimen at 6°C for four consecutive days. Chronic cold stress did not lead to weight differences between Cidec-L^R10G/R10G^ and wildtype Cidec-L^+/+^ mice (Fig. 9A), but Cidec-L^R10G/R10G^ mice exhibited a significant decrease in body temperature, revealing their cold intolerance (Fig. 9B). Post cold exposure, tissue mass of subcutaneous inguinal WAT (iWAT) remained unchanged across genotypes (data not shown), while the classical interscapular BAT from Cidec-L^R10G/R10G^ mutant mice exhibited substantial hypertrophy, approximately doubling in size compared to their Cidec-L^+/+^ counterparts (Fig. 9C). No detectable expression changes in *Ucp1* and other thermogenic genes were observed in either iWAT or BAT (data not shown). However, histological analyses revealed striking morphological alterations in BAT from Cidec-L^R10G/R10G^ mice, including a greater number of medium and large LDs (Fig. 9D). BAT from heterozygous Cidec-L^+/R10G^ mice also showed significantly increased LD size (Fig. 9D) consistent with the observed CIDEC-L haplo-insufficiency in humans. The reduced surface to volume ratio of these large LD makes fat less accessible for lipolysis and thus explains the impaired ability to thermoregulate under cold conditions. In contrast, iWAT of Cidec-L^R10G/R10G^ mice exhibited a morphology comparable to that of Cidec-L^+/+^ mice, without overt changes in LD size (Fig. 9D). Collectively, these findings suggest that the mouse Cidec-L^R10G/R10G^ selectively drives lipid droplet expansion in iBAT, converging with our *in vitr*o molecular data and with the patients’ BAT phenotype.

**Figure 8.**
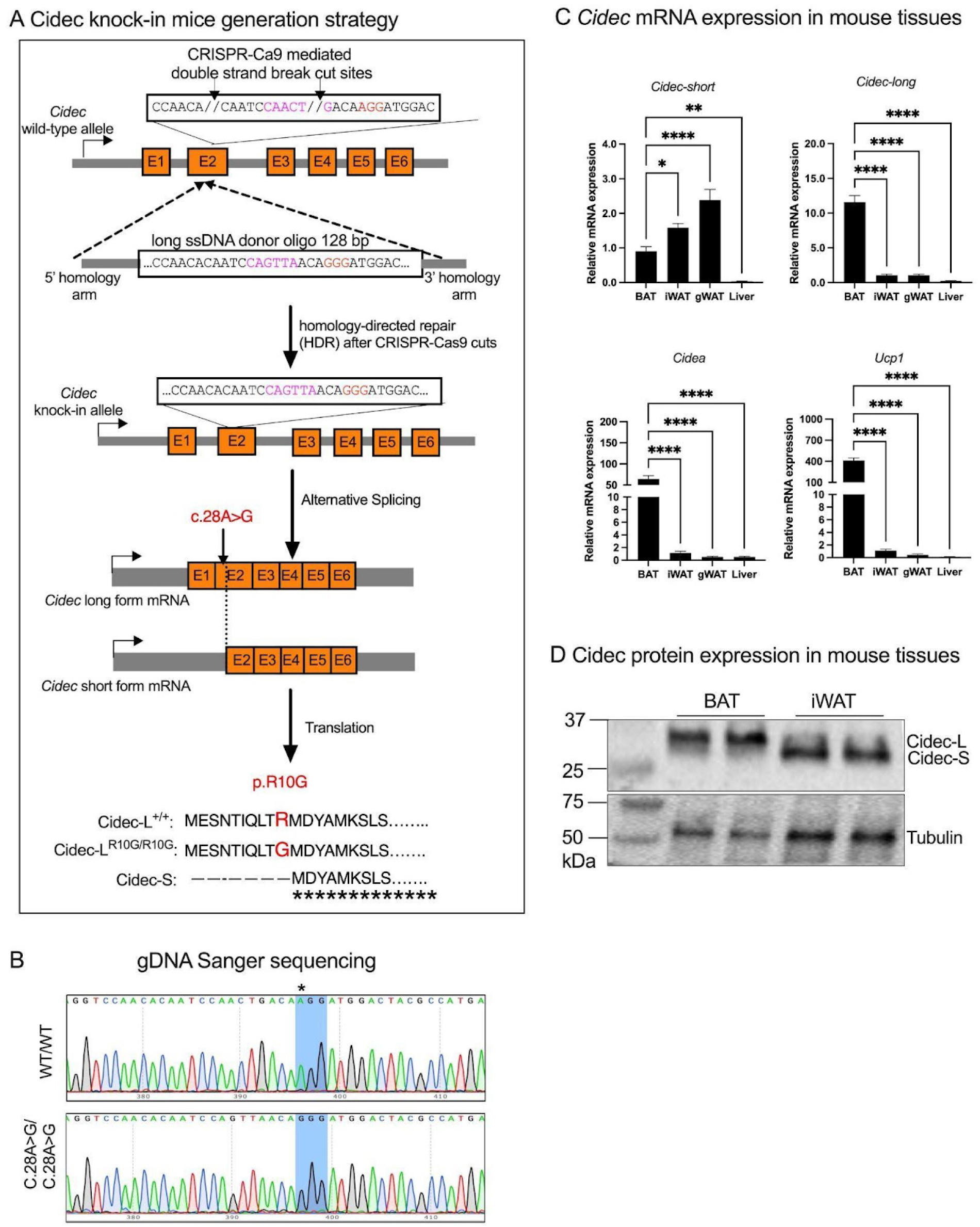
Transgenic knock-in Cidec-L^R10G/R10G^ mouse model generation. **Panel A** shows the schematic of the C57BL/6J mouse model with point mutation (c.28A>G, AGG to GGG, red highlight) at mouse Cidec locus by CRISPR/Cas9-mediated genome engineering. Synonymous mutation (highlighted in Magenta) was introduced to prevent the binding and re-cutting of the sequence by gRNA after homology-directed repair. Like the human mutation, the mouse c.28A>G point mutation selectively alters the Cidec-L protein sequence (p.R10G), without affecting the Cidec-S protein. **Panel B** shows gDNA sequences confirmed by Sanger sequencing. **Panel C** shows mRNA expression of *Cidec-S*, *Cidec-L*, *Ucp1* and *Cidea* gene expression in tissues of iBAT, iWAT, gWAT and liver from 6 weeks old wildtype mice housed at 23 °C. *\*p* < 0.05, *\*\*p* <0.01, *\*\*\*\*p* < 0.0001 were calculated using one-way ANOVA with Dunnett’s multiple comparisons test (n = 5/group). Data are shown as individual measurements and results are presented as means ± SEM. **Panel D** shows immunoblot of endogenous Cidec protein expression in tissues of iBAT and iWAT of 6 weeks old wildtype mice housed at 23 °C, n = 2 mice for each tissue.

**Figure 9.**
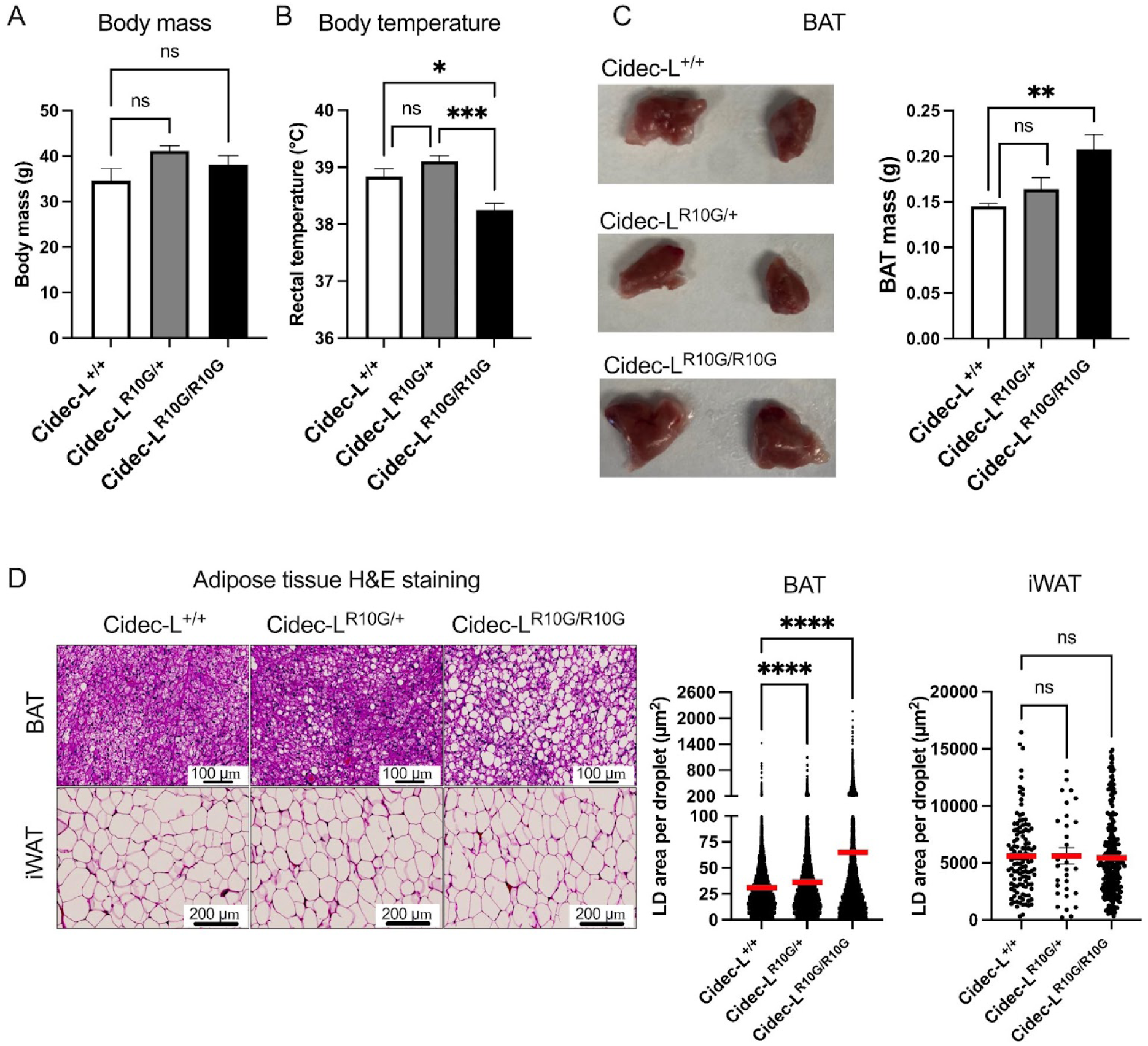
Cidec-L^R10G/R10G^ mutant mice develop enlarged LDs in BAT. **Panel A** shows body weight of male mice fed Western Diet after 4 days of cold exposure at 6°C (n = 6/group). **Panel B** shows the rectal body temperature of male mice fed Western Diet after 4 days of cold exposure at 6°C (n = 6/group). **Panel C** shows representative images and corresponding tissue weight of iBAT from male mice fed Western Diet after 4 days cold exposure (n = 6/group). **Panel D** shows representative Hematoxylin and eosin (H&E) staining of BAT and iWAT, with the corresponding LD size quantifications from the H&E sections. Data are shown as individual measurements and results are presented as means ± SEM. \**p* ≤ 0.05, \*\**p* ≤ 0.01, \*\*\*\**p* ≤ 0.001, \*\*\*\**p* ≤ 0.0001 were calculated by one-way ANOVA.

## Discussion

This study identifies a germline loss-of-function p.R13G variant in human CIDEC-L that segregates in a three-generation family comprising 13 individuals with an obese phenotype characterized by increased subcutaneous fat accumulation, enhanced BAT fat content and insulin resistance (Fig. 1, 2 and Table 1). This hereditary monogenic obesity syndrome related to LD dynamics is distinctive, contrasting with many known familial obesity genes encoding proteins that regulate appetite in the CNS (Ahima et al. 1996; Fan et al. 1997; Huszar et al. 1997; Barsh et al. 2000; Farooqi et al. 2003; Stijnen et al. 2016). It also contrasts with classical familial lipodystrophies associated with LD protein variants which are typically associated with a generalized or partial loss of adipose tissue and severe metabolic disturbances, such as type 2 diabetes liver steatosis, and dyslipidemia. For example, a single human subject with a homozygous CIDEC stop-gained allele (CIDEC-S^E186*^, CIDEC-L^E199*^) was reported with loss of adipose tissues in a partial lipodystrophy phenotype with metabolic syndrome (Rubio-Cabezas et al. 2009). This is mirrored in Cidec null mice under HFD conditions and in genetically obese Cidec null mice (Zhou et al. 2015). This CIDEC-S^E186*^ / CIDEC-L^E199*^ variant would either result in a RNA and protein null allele following nonsense-mediated mRNA decay (NMD) or cause the truncation of both CIDEC proteins, eliminating their binding to LDs, thus causing complete loss of their functions (Rubio-Cabezas et al. 2009). In contrast, the symptomatology of individuals carrying the CIDEC-L^R13G^ variant is marked by an opposite phenotype consisting of progressive subcutaneous visceral, hepatic and pancreatic fat accumulation in the abdomen and proximal extremities, without overt increase in circulating triglycerides or free fatty acids suggestive of some degree of ‘metabolic buffering’ effect offered by an increase in fat expandability (Table 1). Thus, the CIDEC-L^R13G^ variant reported here represents a novel form of heightened adipose tissue depot enlargement accompanied by insufficient increase in fat expandability, resulting in enlarging fat depots and organ fatty infiltration driven by loss of lipid droplet size restriction, which worsens with age.

At the molecular level, our findings on the CIDEC-L^R13G^ variant are aligned with a complex set of functions previously ascribed to CIDE proteins. The discoveries of CIDEC and CIDEA as LD-associated proteins pointed to their role in maintaining the balance between lipid storage in LD and lipolysis (Puri et al. 2007; Puri et al. 2008). This was further supported by the finding that CIDEC directly interacts with the LD binding protein PLIN1, a known modulator of lipolysis (Grahn et al. 2013; Grabner et al. 2021), and can modulate the transcription of *ATGL*, a pivotal lipase in the lipolytic cascade (Singh et al. 2014; Slayton et al. 2019). Additional roles for CIDE proteins include the control of LD size (Puri and Czech 2008; Gong et al. 2011; Xu et al. 2024) by regulating lipid flux between LDs (Gong et al. 2011; Lyu et al. 2021), as well as actions of CIDEA on *UCP1* transcription (Jash et al. 2019).

More recently, Cide proteins were hypothesized to work in a dominant inhibitory fashion, where Cidec-L may block the function of Cidea, which normally enhances LD enlargement (Nordström et al. 2005; Grabner et al. 2021). In line with this notion, we here show that CIDEC-L co-localizes and forms heteromers with CIDEC-S and CIDEA. This aligns with a model where LD size is determined by the ratio of growth-promoting CIDEC-S and CIDEA to growth-restricting CIDEC-L, within CIDE heteromers located at the LD-LD interphase (Nishimoto et al. 2017; Nishimoto and Tamori 2017; Lyu et al. 2021; Ganeva et al. 2023) (Fig. 10). Our phase separation assay implicates the N-terminal structural order in regulating lipid transfer between droplets: the presence of more structured CIDEC-L in a CIDE complex tips the balance towards hydrophilicity, while a higher ratio of disordered CIDEC-S or CIDEC-L^R13G^ tails creates a lipophilic environment at the LDCS (Lyu et al. 2021). This de-repression of CIDEC-S or CIDEA in individuals carrying the CIDEC-L^R13G^ loss-of-function variant therefore manifests itself as adipose tissue enlargement, whereas a complete inactivation of both CIDEC isoforms results in a reverse phenotype characterized as a near-complete atrophy of adipose tissue (Rubio-Cabezas et al. 2009).

**Figure 10.**
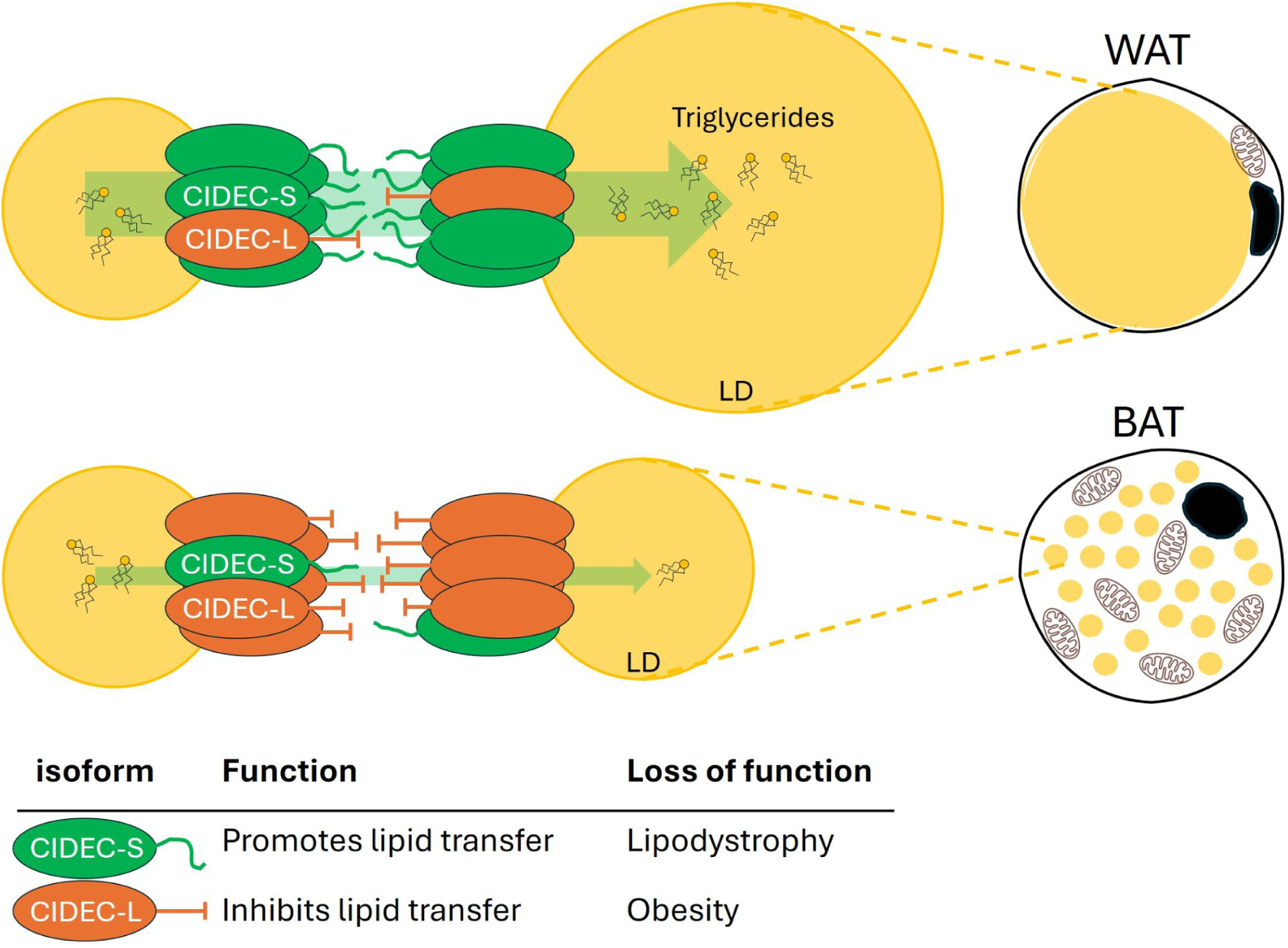
Model depicting the antagonistic effects of CIDEC-L and CIDEC-S on LD dynamics in WAT and BAT. CIDEC-S (green) and CIDEC-L (orange) form heteromers at the lipid droplet (LD) surface. The N-terminal tails protrude into the LD-LD interphase. A higher proportion of disordered CIDEC-S tails promotes condensation, thus facilitating lipid transfer between droplets resulting in unilocular large LD in white adipose tissue (WAT). In contrast, a higher proportion of structured CIDEC-L N-terminal tails prevents condensation and thus restricts lipid transfer between droplets, yielding multilocular and smaller LDs in brown adipose tissue (BAT). A loss of CIDEC-S function results in an inability to efficiently store triglycerides in LD, resulting in a severe lipodystrophy (Rubio-Cabezas et al. 2009). In contrast, loss of CIDEC-L function, as reported here, results in an opposite phenotype of marked obesity due to fat accumulation in BAT and WAT.

Our *in vivo* mouse model presented here provides important mechanistic insights but has inherent limitations in fully recapitulating the human phenotype. While the homozygous Cidec-L^R10G^ knock-in mice exhibited enlarged LDs and adipose tissue hypertrophy in BAT, they failed to develop the pronounced WAT expansion seen in human carriers of the CIDEC-L^R13G^ variant. The relatively high levels of Cidec-L expression in mouse BAT and its low levels of expression in mouse WAT fits with a strong BAT phenotype and lack, or muted, WAT phenotype in homozygous Cidec-L^R10G^ knock-in mice. The exact expression patterns of both CIDEC isoforms remains to be investigated in a wider range of adipose tissue depots in both human and mouse to better quantify such correlations. The discrepancy to the human WAT phenotype could also reflect fundamental species differences in adipose tissue distribution and function which may not be fully captured within the typical lifespan of laboratory mice. Future studies utilizing humanized models or long-term metabolic assessments in aged knock-in mice may provide further insights into the role of CIDEC-L in WAT expansion *in vivo*.

An intriguing question that is raised by these studies is whether the expansion of WAT in humans and BAT in mice in response to CIDEC-L^R13G^ expression is due to increased fat within individual adipocytes (hypertrophy) or to increased numbers of adipocytes in the depots (hyperplasia). The subcutaneous fat biopsy of patient I:1 (Fig. 1C) suggests that the size of the white adipocytes is comparable to control WAT. Thus it may be possible that expression of the CIDEC-L^R13G^ variant in adipocytes or in other adipose tissue cell types causes increases in progenitor cells or in their rates of differentiation. It has been established that crosstalk among adipose cell types is indeed vibrant and important for tissue homeostasis (Kahn et al. 2019; Sakers et al. 2022). Additional experiments on aged Cidec-L^R10G^ knockin mice housed at room temperature will be useful in this regard.

In summary, our findings provide molecular evidence for the dominant-negative effect of CIDEC-L onto CIDEC-S and CIDEA, which function to limit LD size increase. Instead, the described CIDEC-L^R13G^ variant leads to pathological fat accumulation due to unhindered LD growth. This Mendelian disorder of obesity reveals new layers of regulation in lipid droplet dynamics, with potential implications for understanding excessive adiposity and adipocyte lipid storage disorders. This raises the question whether disrupting adipocyte biology by CIDEC-L^R13G^ feeds back to enhancing appetite as well. Future studies will be necessary to determine the long-term metabolic consequences of disrupting LD dynamics and its potential as a therapeutic target to limit cellular fat uptake in patients at risk for obesity. The findings that germline loss-of-function variants in CIDEB may provide protection from liver disease by limiting fat uptake in the liver lends credence to this concept (Verweij et al. 2022).

## Methods

### Oversight

Written informed consent was obtained with the use of consent forms from the Genome Institute of Singapore and the Institute for Human Development and Potential. Studies were carried out under IRB approval from KAUST (23IBEC090) and A*STAR (IRB 2019-087 & DSRB 2022/00719) .

### Variant identification Patients and clinical assessments

#### Sample collection

Genomic DNA samples were collected from saliva of the indicated family members into Oragene Saliva collection vessels (DNAgenotek) and processed for genomic DNA extraction as per manufacturer’s instructions. A skin biopsy was obtained from the affected individual II:4 (Fig. 1A). Written informed consent was received from all patients according to the ethical approvals of the local Institutional Review Board (IRB).

#### Whole exome sequencing

We performed whole exome sequencing of the Trio: affected patient II:4, unaffected mother I:2, and unaffected brother II:8. One microgram of genomic DNA per sample was used for exome capture with Agilent Technologies SureSelectXT All Human ExonV6 Kit. The exome library was prepared on an Ion OneTouch System and sequenced on an Ion Proton instrument (Life Technologies, Carlsbad, CA, USA) using one Ion PI chip. Sequence reads were aligned to the human reference genome (Human GRCh37 (hg19) build) using Torrent Mapping Alignment Program (TMAP) from the Torrent Suite (v5.0.2). The variants were called using the Torrent Variant Caller (TVC) plugin (v5.0.2), and were annotated with the associated gene, location, quality-score, coverage, predicted functional consequences, protein position and amino acid changes, SIFT (Kumar et al. 2009), PolyPhen2 (Adzhubei et al. 2010), Grantham (Grantham 1974) and M-CAP (Jagadeesh et al. 2016) prediction scores, phyloP conservation scores (Pollard et al. 2010) and 5000 genomes Minor Allele Frequencies. Variants were filtered for common SNPs using the NCBI’s “common and no known medical impacts” database (ftp://ftp.ncbi.nlm.nih.gov/pub/clinvar/vcf_GRCh37/) and the Exome Aggregate Consortium (ftp://ftp.broadinstitute.org/pub/ExAC_release/release0.2/). Out of 1306 rare variants (AF<1%), further filtering was conducted to keep only coding, splicing and highly conserved (PhyloP>3) utr/intronic variants that were not detected in the previously 605 in-house sequenced patients, totalling 67 heterozygous and 6 homozygous. Only 5 novel variants, that were not present in public databases and with high deleterious predicted scores (M-CAP>0.15), were kept, including c.-3A>G; p? in the *CIDEC* gene. Sanger sequencing as standard was used to confirm the variants identified and segregation of the phenotype-genotype in the affected individuals. CIDEC-L (NM_001199623) mutation screening was performed with forward primer GGGTTATCTGCTCAGTGTGACC and reverse primer CCCCTGTGACCTGTATACCCAT.

### Clinical assessments in Singapore

#### Study design

This study was conducted over two participants for two test sessions. The first session involved an intravenous glucose tolerance test (IVGTT) to evaluate insulin sensitivity after glucose injection. The second session included whole-body calorimetry to measure energy expenditure, followed with PET/MR imaging to assess brown adipose tissue (BAT) activity. These sessions were scheduled on separate days. Participants were instructed to avoid caffeine, alcohol, and any vigorous physical activity the day before each study session. The present study was conducted according to the guidelines of the Declaration of Helsinki, and all procedures were approved by the Domain-Specific Review Board of National Healthcare Group, Singapore (DSRB approval reference: 2022/00719). Written informed consent was obtained from the two participants before participation.

#### Clinical measurements

Anthropometric measurements included height, weight, neck, chest, waist, upper arm, lower arm, thigh, calf and hip circumference. Body composition such as fat mass, bone mass and lean mass and bone mineral density (BMD) were measured by dual-energy X-ray absorptiometry (DXA) (QDR 4500A, fan-beam densitometer, software version 8.21; Hologic, Waltham, USA).

#### Whole body calorimetry

After fasting for 10-12 hours, participants arrived at the Clinical Nutrition Research Center (CNRC) at 8:00 AM. A registered nurse inserted an indwelling cannula into a forearm vein following a 10-minute rest period, and a baseline blood sample was collected to measure fasting blood parameters. Participants then entered a dual-room whole-body calorimeter (WBC) chamber located at CNRC. These WBC chambers are open-circuit, airtight indirect calorimeters that measure energy expenditure (EE), fat oxidation (FOX), and respiratory exchange ratio (RER) based on oxygen consumption and carbon dioxide production. After a 45-minute resting metabolic rate (RMR) measurement, subjects were exposed to mild cold (approximately 14.5°C) by wearing a cooling vest (Polar Product) inside the WBC chamber for 45 minutes. Blood was collected after participants exited the WBC room.

#### 18F-FDG-PET/MR of brown fat

After exiting the WBC room, participants were taken to the Clinical Imaging Research Center (CIRC) located in the basement of the same building. While still wearing the cooling vest, participants received an intravenous injection of 18 Fluorine Fluorodeoxyglucose (18F-FDG, 3 mCi). Thirty minutes after the injection, PET scans were performed using a hybrid co-registered MR-PET system (Biograph mMR, Siemens Healthcare, Germany) for 60 minutes. An experienced researcher, blinded to the study’s objectives, assessed 18F-FDG uptake in both sides of the neck and supraclavicular regions, excluding bone, muscle, and blood vessels. BAT activity was quantified by calculating the mean SUV. MRI of the supraclavicular fat depot was performed to quantify BAT activity. Image slices with 2 mm slice thickness and in-plane resolution of 2 x 2 mm were acquired using multi-point Dixon sequence (TR=15 ms, 10 echoes, TE1=1.23 ms, ΔTE=1.23 ms) and body matrix coil after anatomical localization. The multi-echo complex data obtained using multi-point Dixon sequence were reconstructed using a multi-scale graph-cut algorithm (Berglund and Skorpil 2017). Multiple regions of interest (ROIs) were drawn within the left and right supraclavicular fat depots on the fat fraction image. A lower threshold of 40% fat fraction was used to exclude the muscle tissue that could have been inadvertently included within the ROI. The proton density fat fraction (PDFF) of BAT was quantified as the mean fat fraction within the ROIs. A separate control PET/MR scan without cold exposure was performed on a separate day.

#### Abdominal fat quantification

Axial images of the abdomen with 3 mm slice thickness, interslice gap of 0.6 mm, and an in-plane resolution of 1.6 × 1.6 mm were acquired using a two-point Dixon sequence (TR = 4.09 ms, TE1 = 1.23 ms, TE2 = 2.46 ms) and body matrix coil. The image slices covering the abdominal region between L1 and L5 vertebrae were acquired during a breath-hold of 18s. A deep learning based automatic segmentation algorithm followed by manual editing was used to segment and quantify the deep subcutaneous (DSAT), superficial subcutaneous (SSAT), and visceral adipose tissue (VAT) compartments (Kway et al. 2021).

#### Liver and pancreatic fat

The liver and pancreatic fat were determined using multi echo Dixon fat-water imaging sequence (TR = 15 ms, 8 echoes, TE1 = 1.23 ms, ΔTE = 1.24 ms) and body matrix coil after anatomical localization. Multiple regions of interest (ROIs) were selected within the liver and pancreas (head-body and tail) carefully excluding the blood vessels and boundaries in the fat fraction image. The liver and pancreatic fat were quantified as the mean proton density fat fraction within the selected ROIs.

#### Skeletal muscle fat

Skeletal muscle fat content from the soleus muscle was determined using magnetic resonance spectroscopy. The muscle spectrum was obtained from a 2 × 2 × 2 cm^3^ voxel within the soleus muscle using point resolved spectroscopy sequence (TR = 2000 ms, TE = 30 ms). The spectrum was quantified using LCModel (Provencher 1993) and the amount of intramyocellular lipids (IMCL) was calculated and expressed as a ratio with respect to water and corrected for T2 losses (Kautzky-Willer et al. 2003).

#### Intravenous glucose tolerance test (IVGTT)

The insulin-modified minimal model IVGTT protocol was adopted for the study. Intravenous catheters were inserted on opposite arms of participants for blood sampling and intravenous infusion. After baseline blood samples were collected, an IV bolus of 50% dextrose was administered over a duration of 1 minute. The amount of dextrose infused was according to the dosing amount of 300 mg/kg body weight. For the first 10 minutes after bolus administration, blood samples were collected at the following time points: 2, 3, 4, 5, 6, 8 and 10 minutes. This initial collection is required for the assessment of first phase insulin secretion. The test continued with blood sampled at frequent time intervals over 3 hours: 14, 19, 22, 25, 30, 40, 50, 70, 100, 140 and 180 minutes. If the study participants are resistant to insulin, the glucose concentration decay will be slow. To overcome this, the insulin-modified protocol includes an insulin infusion at 20 minutes after the glucose bolus at a standard dose of 0.03 U/kg body weight. The MINMOD Millennium software was used to analyse the IVGTT data using the Bergman minimal model. The computer program provides information on several parameters using glucose and insulin values obtained during the test. This information includes the acute insulin response to glucose (AIRg), disposition index (DI), insulin sensitivity (SI), glucose effectiveness (Sg) and insulin resistance (IR). Blood glucose was measured using an on-site glucose analyzer (YSI 2300 STATPLUS; YSI Incorporated, Life Sciences, Yellow Springs, OH, USA). Serum insulin was measured by NUH Referral Laboratory, NRL (licensed by Licensing & Accreditation, Health Regulation Division, Ministry of Health, Singapore) using Abbott Alinity chemiluminescent immunoassay protocol. IVGTT was conducted in the Investigational Medicine Unit (IMU), National University of Singapore.

#### Blood analysis

Blood samples were submitted to the National University Hospital (NUH) referral laboratory for biochemical assessment of renal, liver, and thyroid function. Venous blood samples were collected during fasting and 1-hour mild cold exposure sessions (WBC) to measure plasma concentrations of glucose, insulin, total cholesterol, LDL, HDL, and triglycerides at NUH. Plasma adiponectin, CRP, FGF-21, ghrelin, GLP-1, IL-6, leptin, irisin, and apelin were analyzed using the MILLIPLEX MAP Human Metabolic Hormone Magnetic Bead Panel (Millipore) on the A*STAR SIgN Multiplex Analysis of Proteins (MAP) platform. Free fatty acid concentrations were determined using a colorimetric assay (Abcam).

### Patient-derived white adipocytes

#### Fibroblast collection

Primary cutaneous dermal fibroblasts were derived from a skin biopsy of individual II:4. Briefly, biopsy was incubated in trypsin overnight at 4°C to enable the peeling of the epidermis from the dermal compartment. Dermis was chopped up and stuck to a 10 cm plastic dish allowing the fibroblasts to migrate out of the dermal fragments. Fibroblasts were cultured in DMEM supplemented with 10% fetal bovine serum and 2 mM L-glutamine. Control primary fibroblasts used as WT controls were obtained from the SRIS Asian Skin Biobank with informed consent and prior IRB approval.

#### Induced pluripotent stem cell (iPSC) generation

Primary cutaneous fibroblasts were reprogrammed using the CytoTune™-iPS 2.0 Sendai Reprogramming Kit (Thermo Fisher Scientific, A16517) in accordance with manufacturer’s instructions. Briefly, fibroblasts were transduced and after 7 days, were plated onto Matrigel Basement Membrane Matrix (Corning, 354234) in mTeSR1 medium (STEMCELL Technologies, 85850). iPSC colonies were picked between days 17-28 and maintained on Matrigel and mTeSR1 for expansion.

#### Differentiation into white adipocytes

iPSC were differentiated into mesenchymal progenitor cells (MPC) and subsequently matured into white adipocytes as previously described in Lee *et al*. (Lee and Cowan 2014).

#### LD size quantification

For Oil Red O staining, cells were fixed in 10% formalin for 10 minutes at room temperature and washed twice with PBS. After two washes with 60% isopropanol, fixed cells were stained with 0.15% Oil Red O in 60% isopropanol for one hour. After two brief washes in 60% isopropanol, cells were kept in PBS at 4C until imaging on an Olympus brightfield microscope with 40x magnification. For immunofluorescence, cells were grown on a Nunc^TM^ Lab-Tek^TM^ 4-well glass chamber slides and fixed for 15 min in ice cold 4% (w/v) paraformaldehyde. Permeabilization using 0.6% (v/v) Triton-X in PBS was performed for 15 min, prior to incubation with LipidTOX Deep Red (H34477, Thermo Fisher) for 1 hour at 4C. Counter staining of nuclei was performed using 1 ug/ml DAPI for 20 minutes and cells stored at 4C in PBS. Images were acquired on an inverted Zeiss LSM 800 confocal microscope through a 40x immersion oil lens. LD size was quantified using ImageJ by thresholding and running the particle analyzer. Parameters and statistical tests are indicated in the respective figure legends.

#### qPCR

Total RNA was isolated from cells using the RNeasy Mini Kit (Qiagen). A total of 1 µg of RNA was reverse transcribed with the Iscript^TM^ cDNA Synthesis Kit (Bio-Rad), and transcript levels were determined with SYBR Green on the QuantStudio 5 Real-Time PCR System. Primers are indicated in Table 2. Repetitions, reference genes and statistical tests are indicated in the respective figure legends.

**Table 2.**
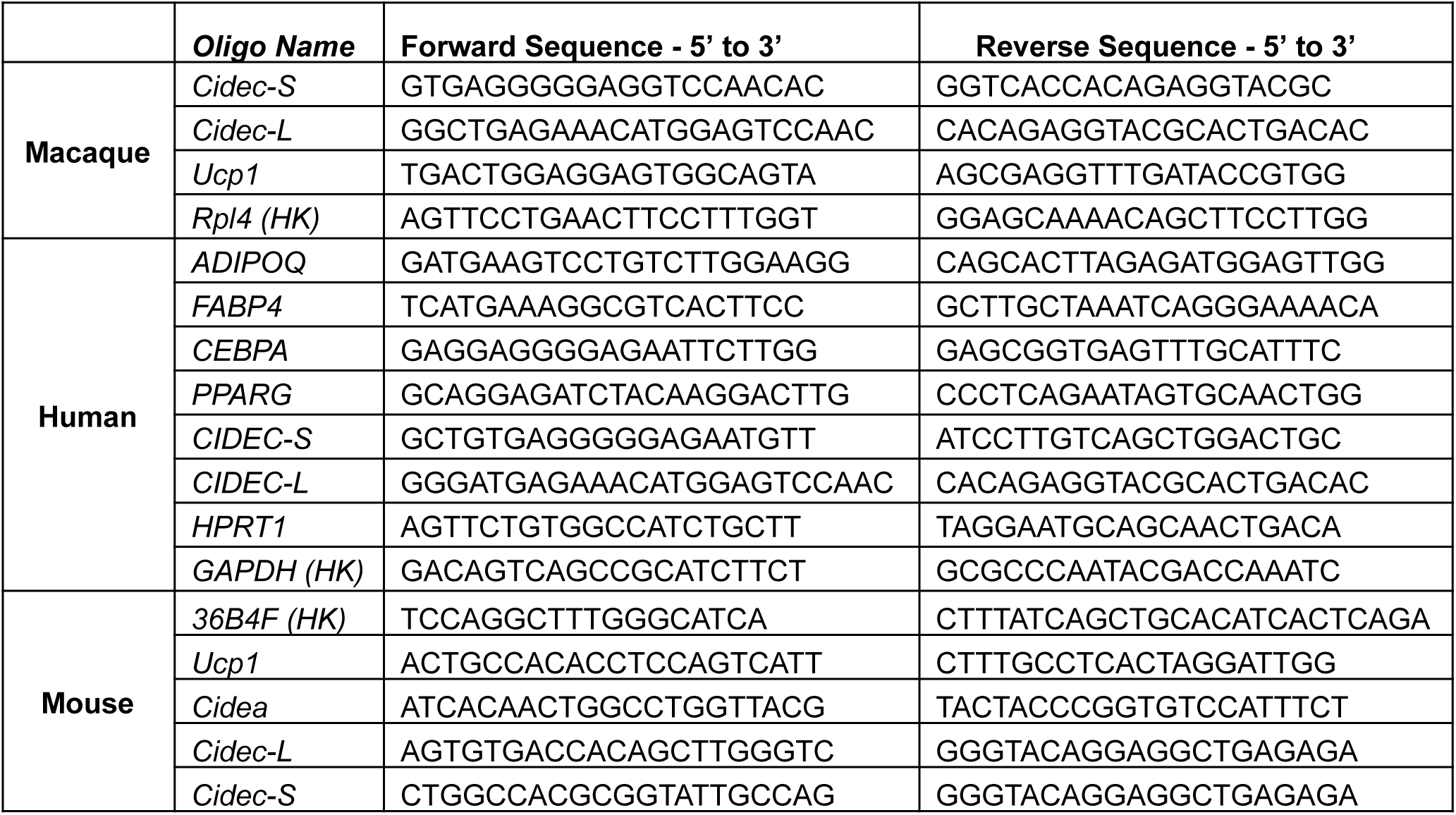
List of qPCR primers.

### Mechanistic studies

#### Plasmid construction and mutagenesis

Full-length cDNAs encoding human CICEC-S, CIDEC-L, CIDEA and PLIN1 were cloned from cDNA of human white adipocytes and subcloned into pCMV5-HA, pCMV5-Flag, pEGFPN1, or pEGFPC1 vectors. PCR amplified full-length CIDEC was subcloned into XhoI–EcoRI sites of pCDNA-3.1(–) and pEGFPN1 vectors, or NdeI–BamHI sites of pCMV5-HA and pCMV5-Flag vectors. Amino acid substitutions in CIDEC-L were generated by PCR site-directed mutagenesis from wild-type CIDEC-L-GFP. The fidelity of all plasmid DNA constructs was verified by sequencing. Constructs for *in vitro* protein expression were cloned by insert coding sequence of CIDEC (aa 1-135) into a modified pET11 expression vector: a His6 -tag followed by a solubility tag, MBP. A TEV cleavage site and a GFP tag were located at the N terminus and C terminus of CIDEC (aa 1-135), respectively. The fusion protein was expressed and purified from bacteria.

#### Cell culture, treatment, and transfection

HEK293T cells and 3T3-L1 preadipocytes were cultured in DMEM (Invitrogen, USA) supplemented with 10% FBS (Invitrogen, USA), 2 mM L-glutamine, 100 U/ml penicillin, and 100 µg/ml streptomycin. Cells were incubated at 37°C in a humidified incubator containing 5% CO_2_. To promote the formation of LDs, cells were treated with 200 µM oleic acid (OA; Sigma, USA) conjugated to fatty acid–free BSA at a molar ratio of 6:1 and cultured for 16 h. HEK293T cells were transfected with plasmids using Lipofectamine 2000 according to the manufacturer’s instruction (Invitrogen). Electroporation of plasmid DNAs into 3T3-L1 preadipocytes was performed using Amaxa Nucleofector II (Lonza), program A-033, according to the manufacturer’s instruction.

#### LDs size analysis and statistics

Cells were cultured at 37℃, 5% CO_2_, and 80% humidity. After loading with 200 µM OA for 16 h, cells were washed 3 times by PBS, and then fixed with 4% PFA. LDs were stained with 1:1000 diluted Bodipy 493/503 (D3922, Invitrogen) or LipidTOX Deep Red (H34477, Invitrogen) for 20 minutes. Nucleus were stained with 1:10,000 diluted Hoechst 33342 (1 mg/ml, 62249, Invitrogen) for 20 minutes. Images were acquired on a Zeiss LSM 900 with Airyscan 2 confocal microscope with 100X oil immersion objective and 405/488/561/640 laser. Manual measurement of LDs size was performed using ImageJ (NIH), and Imaris 8.0 (Bitplanet). LDs stained with fluorescent dyes were detected by default Threshold (ImageJ) and surface using background subtraction method (Imaris), and the numbers, sizes, and volumes of LDs were automatically measured.

#### FRAP-based lipid exchange rate assay

Lipid exchange rate assay was performed as previously described (Gong et al. 2011; Wang et al. 2019) with some modification . 3T3-L1 preadipocytes co-transfected with indicated plasmids (PLIN1 and CIDEC-S, CIDEC-L or CIDEC-L^R13G^) were incubated with 200 µM oleic acid and 1 mg/ml BODIPY 558/568 C12 fatty acid (Molecular Probes, USA) for 18h and transferred to fresh medium 1h before FRAP experiments. Live cells were visualized under a confocal microscope (A1Rsi, Nikon, Japan) using a 100 X oil-immersion objective. LD pairs in a range of 1-6 μm in diameter with clear green fluorescent signal at lipid droplet cell surface (LDCS) in 3T3-L1 preadipocytes were photobleached, whereas LD pairs in adipocytes were randomly selected. At least 70% of LD total area were photobleached for 1 s at 100% laser power (561 solid state laser), followed by time-lapse scanning with a 1-s interval for pre-adipocytes. The same photobleaching process was repeated three times. Mean optical intensities (MOI) within LD core regions were measured simultaneously. Unbefitting data were filtered out based on the criteria in the lipid exchange rate assay. Digital detectors gain and laser power were set to avoid overexposure and ensure accurate quantification of fluorescence.

#### Co-immunoprecipitation

Co-immunoprecipitation was performed according to previous procedure (Ye et al. 2009). Cells transfected with the indicated plasmids were washed with PBS and then lysed in IP buffer (20 mM Tris-HCl, pH 7.4, 150 mM NaCl, 1% Triton X-100, 1 mM EDTA, 1 mM EGTA, cocktail (Roche)) by sonication. M2 beads crosslinked with Flag antibody were used for immunoprecipitation. The resulting protein complexes were then subjected to Western blot analysis with the indicated antibodies. Antibodies against Flag (Sigma, F1804, 1:1000, USA), and HA (Santa Cruz Biotechnology, sc-7392, 1:1000, USA) were commercially acquired. The blots were detected using HRP-conjugated secondary antibodies (GE Health, UK) and the ECL-plus system. Membranes were blocked with 5% fat-free milk.

#### Structural modelling

Protein structures were modelled using AlphaFold3 with standard parameters on the AlphaFold server (Abramson et al. 2024). Either monomers or hexamers, based on findings in Choi *et al*. (Choi et al. 2017), were modelled with residues and confidence indicated in the figure legends. Structures for each molecule were modelled at least ten times.

#### Protein purification

Recombinant CIDEC and mutants were expressed in BL21 (DE3) cells. Bacteria were grown in suspension at 37°C to an optical density of 0.6 and protein expression was induced by adding 1 mM isopropyl beta-d-thiogalactopyranoside (IPTG). Cells were induced overnight at 18°C and collected by centrifugation and lysed by sonication in lysis buffer (25 mM Tris-HCl, pH 7.4, 1 M NaCl, 5% glycerol, protease inhibitor cocktail (Complete, Roche), 5 mM βME). Lysates were cleared by centrifugation 20,000 *g*, 60 min, 4℃ (JA-25.50 rotor, Beckman Coulter). Centrifuge-cleared lysate was applied to Ni-NTA agarose (Life technologies), washed with lysis buffer containing 20 mM imidazole, and eluted with the same buffer containing 300 mM imidazole and 2 mM βME.

#### Cell free phase separation assay

Proteins were pre-cleared via high-speed centrifugation. The concentration was determined by measuring the absorbance at 280 nm using a NanoDrop spectrophotometer (Thermo Scientific) before cleavage. *In vitro* phase separation assay was performed in the same buffer containing 25 mM Tri-HCl, pH 7.4, 150 mM NaCl, and 10% PEG 4000. MBP tag were cleaved before droplet assembly with TEV protease. Droplets were assembled in 384 multi-well microscopy plates (384-well microscopy plates, PerkinElmer), covered with optically clear adhesive film and observed under a Zeiss LSM 900 microscope equipped with 60’ and 100’ oil immersion objectives.

### Animal Work

#### Animal Husbandry

All animal work post generation was performed at the University of Massachusetts Chan Medical School in accordance with the Institutional Animal Care and Use Committee.

#### Mouse generation

*Cidec* point mutation knock-in mice were generated on a C57Bl/6J genetic background by Cyagen Biosciences (USA) and kept on a C57Bl/6J genetic background. To do so, the gDNA to mouse *Cidec* gene, the donor oligo containing the p.R10G (AGG to GGG) mutation and two synonymous mutations p.Q7= (CAA to CAG) and p.L8= (CTG to TTA), Cas9 were co-injected into fertilized mouse eggs to generate targeted knock-in offspring (Fig. 8A). The two synonymous mutations were introduced to prevent the binding and re-cutting of the sequence by gRNA after homology-directed repair. F0 founder animals were identified by PCR followed by sequence analysis, which were bred to wildtype mice to test germline transmission and F1 animal generation. Full details regarding the mutant mice generation, genome DNA genotyping is provided in Figure 8.

#### Mouse genotyping

Genome DNA were isolated from ear samples of 21 days old mice, then samples were digested with 100 μl tail lysis buffer (50 mM Tris pH 8.0, 50 mM Kcl, 2.5 mM EDTA, 0.45% NP40, 0.45% Tween 20, 100 μg/ml proteinase K), 65 ^°^C overnight, 95 ^°^C 10 min to inactivate proteinase K. Genotyping was performed by genome PCR analysis using following oligonucleotide primers, forward primer 5’-CACAGCTTGGGTCGGAGAAA-3’, reverse primer 5’-CTGCCCGTATCTGTGTTTCCA-3’. PCR were carried out in a 20 μL volume system using KAPA HiFi HotStart ReadyMix (Roche, 07961316001), 10 μM primers, and the above gDNA template. The PCR products were then used for confirmation by Sanger sequencing.

#### Western diet feeding

At 16-week-old, all mice were fed with Western diet for another 16 weeks (Inotiv, TD.88137, 42% from fat, 0.2% total cholesterol, high in saturated fatty acid and higher in sucrose).

#### Cold exposure

After 16 weeks Western diet feeding, mice were singly housed at 6 °C for four days, with free access to Western Diet pellets and water. Body weight and body temperature were monitored, the latter of which was measured using a rectal probe (Braintree Scientific, RET-3).

#### qPCR of mouse samples

Total RNA was isolated from mouse tissues using Trizol Lysis Reagent (QIAGEN) following the manufacturer’s instructions as previously described. Mouse 36B4 served as controls for normalization. Primer sequences used for qRT-PCR analyses are listed in Table 2.

#### Western Blot of mouse samples

Tissues were homogenized in RIPA buffer (50 mmol/L Tris-HCl (pH 8.0), 5 mmol/L EDTA, 0.25 mol/L NaCl, with 1x Halt protease inhibitors (Thermo Scientific, 78430). Tissue lysates were then quantitated by bicinchoninic acid (BCA). Protein lysates were prepared for gel electrophoresis in 1x Laemmli buffer with B-mercaptoethanol and heated for 10 min at 95°C. Proteins were resolved by SDS-PAGE using 4-15% Mini-PROTEAN TGX gels, followed by transferring to nitrocellulose. Nitrocellulose membranes were blocked in 5% Non-fat milk in 0.1% TBS-T (20mM Tris pH7.5, 150mM NaCl and 0.1% Tween-20) for 1 hour at room temperature The following antibodies (Ab) were used: anti-Cidec Ab (Thermo Fisher, PA5-30793, 1:1000 dilution), anti-Tubulin Ab (Sigma-Aldrich, sc5286, 1:1000 dilution). Immunoblotting was performed by incubating membranes overnight at 4°C, on a rolling shaker in antibodies, followed by washing with 0.1%TBS-T. Secondary HRP Antibodies were diluted 1:10,000 in 0.1% TBS-T with 5%wt/v bovine serum albumin and incubated for 45min-1hr at room temperature, with shaking. Membranes were washed with TBS-T, followed by enhanced chemiluminescence (ECL) (Perkin Elmer, NEL104001EA).

#### Histological analysis of mouse adipose tissues

For immunohistochemistry, tissue samples were fixed in 10% Formalin and embedded in paraffin. Sectioned slides were then stained with H&E, at the UMass Chan Medical School Morphology Core. Photos from the H&E staining were taken with an Axiovert 35 Zeiss microscope (Zeiss) equipped with an Axiocam CCl camera at indicated magnification. LD sizes from the BAT and WAT tissues were quantified with ImageJ. An ImageJ macro language program was written to analyze each H&E staining image using ImageJ-Labkit plugin to classify pixels as either lipid or background.

#### Statistical analysis

Data were analyzed in GraphPad Prism 8 (GraphPad Software, Inc.). Statistical tests used and p-value cutoffs are indicated in the respective figure legends. All comparisons between two groups were performed with student unpaired two-tailed Student’s t-test and One-way ANOVA with the following demonstration of p values in the panels: * *p* <0.05, ** *p* <0.01, *** *p* <0.001, **** *p* <0.0001. For experiments with a two-factorial design, multiple comparisons were analyzed by two-way ANOVA to determine the statistical significance between groups based on one variable. The data are presented as means ± SEM or means ± SD as indicated in figure legends

## Data Availability

All data produced in the present study are available upon reasonable request to the authors

## Acknowledgements

We thank all members of our respective laboratories for useful discussions of the data during development of the project. F. P. is a recipient of a long-term European Molecular Biology Organization (EMBO) postdoc fellowship and a short-term EMBO travel fellowship. Her research is supported by the Singapore Ministry of Health’s National Medical Research Council under its Young Individual Research Grant scheme (Project ID MOH-000549-01) and A*STAR under its Career Development Award (Project number C210112002). L. J. T. is supported by the A*STAR Career Development Fund (CDF C243512024). We thank Prof. Patrick TAN and the Genome Institute of Singapore (GIS) for supporting the costs associated with the travel and clinical phenotyping of two patients in Singapore. Research in L.P.’s laboratory was supported by the National Key R&D Program of China (2024YFA1802802) and the National Natural Science Foundation of China (92357302). Funding by the National Institutes of Health (DK130852 and DK116056 to M.P.C) and by the Isadore and Fannie Foxman endowed Chair in Medical Research at the University of Massachusetts Chan Medical School to M.P.C. is gratefully acknowledged. (SR/MED/GENT/16/01). B.R. is a fellow of the Branco Weiss Foundation (Switzerland) and an EMBO Young Investigator (Europe). The research reported in this publication was supported by funding from King Abdullah University of Science and Technology (KAUST) and from GIS at A*STAR (Singapore).

## Author Contributions

F.P., H.W., J.Q.W., L.X., P. L., M.P.C., B.R., designed the study and interpreted the data. S. T., A. Y. J. N., and V.B. performed whole exome sequencing. L.S., H. J. G., S. A. S., S. S. V., P. S. L., K. A. T. E.-S., P. P. A.-W., E. P.-P., E. M. C. C.-D. L. P. and M. K. S. L. designed and performed the patient experiments as well as collected patient data. F.P., J.W., L.J.T, C. Y. C. and G. N. performed and analyzed the *in vitro* experiments. H. W., M. K, L. M. L. and S. N. performed and analyzed the mouse experiments. C. B., Q. L., M. K. S. L., L. X., P. L. M. P. C. and B. R. provided oversight over this study. F.P., H.W., M.P.C. and B.R. wrote the manuscript. All authors reviewed the manuscript for important intellectual content and agreed to the decision to submit the manuscript for publication.

## Competing interests

None declared.

## Notes

### Competing Interest Statement

The authors have declared no competing interest.

### Author Declarations

Institutional Review Boards of the Agency for Science, Technology and Research in Singapore and King Abdullah University of Science and Technology in Saudi Arabia; as well as the Domain Specific Review Board of the Singapore National Healthcare Group gave ethical approval for this work

